# Biallelic germline variants in the hematologic malignancy predisposition gene *DDX41* cause retinal dystrophy through dysregulation of retinal homeostasis

**DOI:** 10.64898/2026.01.28.26344834

**Authors:** Zoéline Mars, Andrea Zanetti, Karolina Kaminska, Takero Miyagawa, Duanya Liu, Aline Antonio, Gavin Arno, Isabelle Audo, Carmen Ayuso, Hafiz Muhammad Jafar Hussain, Xuan Bao, Pilar Barberán-Martínez, Béatrice Bocquet, Anna Boguszewska-Chachulska, Christel Condroyer, Pierre David, Hélène Dollfus, Lucas Fares-Taie, Lidia Fernández-Caballero, Gema García-García, Victor Michel, Chiara Ida Guerrera, Vincent Jung, Line Kessel, Louise Gioja, Siying Lin, Ewa Matczynska, Jose M. Millán, Abigail R. Moye, M Pilar Martín-Gutiérrez, Mathieu Quinodoz, Matthieu P. Robert, Jerome E. Roger, Rui Sousa-Luis, Saoud Tahsin Swafiri, Slawomir Teper, Isabelle Meunier, Olivier Patat, Mark E. Pennesi, Karin A.W. Wadt, Meng Wang, Andrew R. Webster, Paul Yang, Li Yumei, Christina Zeitz, Frederic Rieux-Laucat, Stéphane Giraudier, Rui Chen, Sebastian M. Fica, Carlo Rivolta, Marie Sebert, Jean-Michel Rozet, Isabelle Perrault

## Abstract

Leber congenital amaurosis (LCA) and Early-onset severe retinal dystrophy (EOSRD) manifest within the first months and the first years of life, respectively. They are the leading cause of severe vision impairment in childhood. Using next generation sequencing, we identified eight families of patients with LCA/EOSRD carrying biallelic combination of six germline variants in *DDX41*, encoding a DEAD-box ATPase RNA helicase involved in RNA splicing, innate immunity and hematopoiesis. In fibroblasts from a patient carrying the homozygous missense variant c.1187T>C (p. Ile396Thr) and in the retina of *Ddx41^I396T/I396T^* mice, DDX41 protein expression was decreased. Electroretinogram recordings in these animals also revealed significant visual dysfunction since the first month of age, supporting a pathogenic role of DDX41 in retinal physiology. Immunohistochemical staining showed that the protein localized to nuclei in all major retinal cell types and to photoreceptor synapses, while biochemical assays showed that LCA/EOSRD variants disrupt DDX41 interactions with RNA through misfolding or the formation of non-productive aggregates, resulting in loss-of-function. Transcriptomic profiling of mutant mouse retinas revealed dysregulation of gene networks associated with Müller cells (MCs), glial cells essential for maintaining retinal structure, metabolic balance, and immune surveillance. The dysregulated pathways chiefly involved cell morphogenesis and junction formation, consistent with immunohistological analyses of widespread architectural disruption and nuclear disorganization, identifying MCs as a site of dysfunction. Together, these findings establish for the first time the involvement of *DDX41* in LCA/EOSRD and provide new insights into the role of helicases in retinal homeostasis.

## INTRODUCTION

Helicases are essential enzymes that unwind nucleic acid structures, playing critical roles in both DNA and RNA metabolism^1,2^. Among these, the Asp-Glu-Ala-Asp (DEAD)-box ATPase DDX41 is a notable member, with functions spanning RNA metabolism, genomic integrity, and innate immunity. In the nucleus, DDX41 regulates RNA splicing^3^, processes small nucleolar RNA (snoRNA) and ribosomal RNA (rRNA)^4–6^, resolves R-loops, RNA-DNA hybrids which are crucial for transcription regulation, and participates in DNA repair^7,8^. In the cytoplasm, DDX41 functions as a sensor for dsDNA from infections or cellular damage, activating a cGAS-mediated type I interferon response^9^.

Monoallelic pathogenic germline *DDX41* variants cause the most frequent inherited susceptibility to adult-onset myeloid malignancies, including myelodysplastic syndrome (MDS) and acute myeloid leukemia (AML) with a median age of disease onset of 69 years^3,10^. In most patients, hematopoietic progenitor cells acquire second somatic *DDX41* variants, most frequently the p.R525H hotspot, in the second allele during leukemogenesis^11,12^. Some individuals also develop granulomatous or immune disorders, including sarcoidosis, systemic lupus erythematosus, asthma, eczema, or juvenile arthritis, in the absence of malignancy^13,14^. The mechanisms linking DDX41 deficiency to myeloid neoplasms and immune dysregulation remain incompletely understood, though R-loop accumulation in human and animal models is thought to drive genomic instability, cGAS-STING-mediated inflammation, and disrupted snoRNA processing^12^.

In contrast to these adult-onset phenotypes, biallelic germline missense *DDX41* variants have been reported in rare Mendelian disorders. To date, three individuals have been described. Two siblings presented with mild dysmorphic features, psychomotor delay, and intellectual disability (NM_016222.4:c.962C>T, p.P321L and c.937G>C, p.G313R)^15^, with one developing blastic plasmacytoid dendritic cell neoplasm (BPDCN) in the first decade. A third patient exhibited bone dysplasia, ichthyosis, and dysmorphic features(c.465G>A, p.M155I and c.1033G>A, p.G345L)^16^. None were reported to initially display ocular phenotypes consistent with retinal dystrophy.

Inherited retinal diseases (IRDs) are clinically, genetically and pathophysiologically diverse neurodegenerative disorders affecting photoreceptors. Leber congenital amaurosis (LCA) and early-onset severe retinal dystrophy (EOSRD) are the earliest and most severe forms, presenting in the first months and first years of life, respectively, and together represent a leading cause of childhood blindness, affecting roughly 20% of children in schools for the blind. Investigating the molecular basis of LCA/EOSRD, we identified six unique germline *DDX41* variants in biallelic states (homozygous or compound heterozygous) in eleven patients from eight families. In some patients, the retinal disease was accompanied by neurological or skeletal anomalies.

Among these, a recurrent DEAD domain variant (c.1187T>C; p.I396T), identified in five families, was analyzed using patient-derived fibroblasts and by a homozygous knock-in mouse model. Functional studies revealed reduced DDX41 protein levels in both patient fibroblasts and mouse retinal tissue. Although overt retinal degeneration occurred later, abnormal ERG responses were identified as early as one month. RNAseq and immunofluorescence analyses showed disrupted Müller cell gene networks and mislocalized cell bodies, seemingly indicating glial dysfunction and retinal disorganization. These findings expand the spectrum of DDX41-related disorders and reveal a distinct developmental phenotype in the absence of neoplastic abnormalities.

## RESULTS

### Biallelic *DDX41* Variants are Identified in Unresolved LCA/EOSRD Cases

Whole exome sequencing (WES) initially identified three individuals with EOSRD (P1-P3; **Fig. 1a**) from two families carrying variants in *DDX41* (NM_016222.4). The two siblings from the first family (P1-P2) were homozygous for c.1187T>C (p.I396T, M1), whereas the proband from the second family (P3) carried this variant in trans with c.1015C>T, as demonstrated by parental segregation analysis (p.R339C, M2). Collaboration expanded the cohort to eight additional patients (P4-P11) from six unrelated families. Additionally, analysis of a previously identified family^15^ (P12-P13) were included to assess retinal phenotypes bringing the total to thirteen affected individuals from nine families across six countries (**Fig. 1a**). Variants comprised homozygous or compound heterozygous missense and frameshift changes, with the M1 and M2 alleles observed in additional families. Specifically, M1 was homozygous in P4-P6 and in compound heterozygosity with a private frameshift deletion c.305_306del (p.K102Rfs*32, M3) in P7, whereas M2 was homozygous in P8-P9. Other patients carried private variants: P10 was compound heterozygous for a variant affecting the last nucleotide of exon 8 c.798G>A (p.Ser266=, M4) and a frameshift c.418_419insGTAG (p.Asp140Glyfs*19, M5). Minigene assay analysis showed that the variant resulted in skipping of exon 8 p.Leu216Glyfs*37 (**Supplementary Fig. 1**). P11 was homozygous for c.655C>T (p.R219C, M6). Previously reported Danish siblings (P12-P13) were compound heterozygous for c.962C>T (p.P321L, M7) and c.937G>C (p.G313R, M8)^15^.

**Fig. 1.**
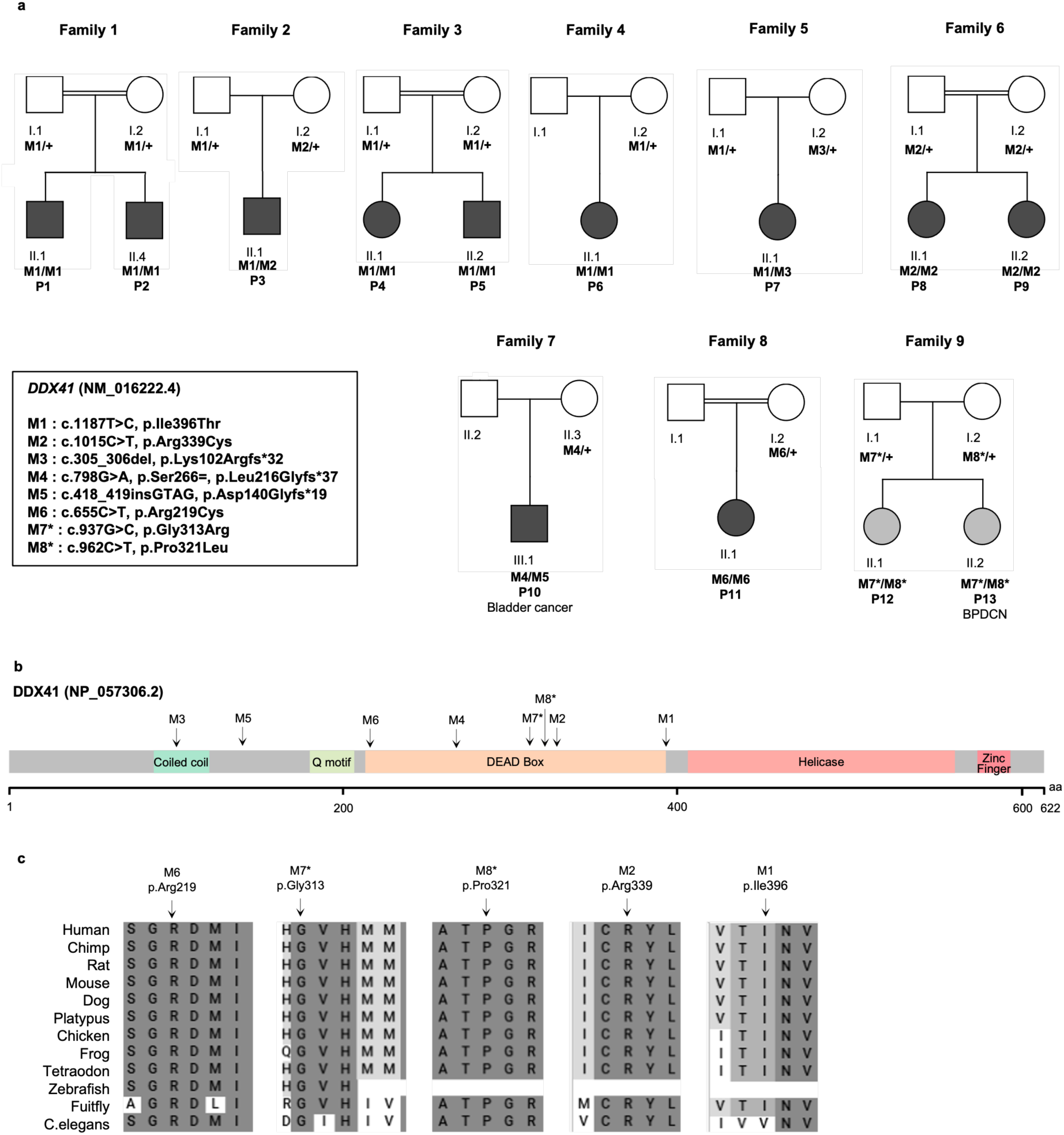
Pedigrees, *DDX41* variant segregation, protein domain organization, variant distribution, and conservation across species. (a) Pedigrees of the nine analyzed families, showing DDX41 variants (M1-M8) described at the cDNA and protein levels, and their segregation. “+” indicates the normal allele; asterisks (*) mark variants previously reported in Diness *et al.* 2018^15^ (Family 9). Symbols filled in black denote individuals affected with EOSRD, with or without extraocular involvement (**Supplementary Table 2**); those filled in grey indicate visual symptoms without complete ophthalmological evaluation (Family 9). (b) Protein domain organization and positions of DDX41 variants. Variants are predominantly located in the DEAD-box domain. aa, amino acids. (c) Conservation of the amino acid sequence at the variant sites across species. The arrow indicates the amino acid affected by the variant. BPDCN: blastic plasmacytoid dendritic cell neoplasm.

According to American College of Medical Genetics and Genomics (ACMG) criteria, the frameshift variant M3 was classified as pathogenic, whereas M1 and M5 were classified as likely pathogenic; M2, M4, M6, M7, and M8 were considered variants of uncertain significance. All variants were ultra-rare (minor allele frequency <1×10⁻⁵) or absent from gnomAD v2.1.1, with no homozygotes reported. The affected residues are highly conserved and cluster within the DEAD-box helicase domain, which mediates ATP binding, hydrolysis, and RNA interaction^1^ (**Fig. 1b-c and Supplementary Table 1**). Notably, all variants except M4 have previously been implicated in acute myeloid leukemia or familial myelodysplastic syndromes^17^. No additional biallelic *DDX41* variants were identified in an independent cohort of 420 unresolved retinal dystrophy cases, including rod-cone dystrophy (RCD) and cone-rod dystrophy (CRD), using a combination of gene panel analysis and Sanger sequencing.

### Biallelic *DDX41* Variants Cause Retinal Dystrophy with Variable Neurological and Skeletal Features

Clinical features of the thirteen affected individuals (5M:8F) are summarized in Supplementary Table 2. Retinal and neurological symptoms were noted in all families except 2 and 3, which showed retinal involvement only. It is noteworthy that retinal involvement in some patients (Families 1, 2, and 5) was reported following an infectious episode. Currently, no patient presents with hematologic symptoms, and available hematologic assessments showed normal blood counts.

### *DDX41* Variants Alter DDX41 Stability and RNA Interaction

DDX41 is predicted to bind RNA on a concave interface formed by the HELIC and DEAD domains, whose stable interaction with RNA is promoted by ATP binding^18^ (**Fig. 2a**). Intriguingly, a Zn^2+^ finger domain is positioned to cap the RNA binding interface and may modulate DDX41 activity. Although the specific role of DDX41 in the nucleus and its target interacting RNAs remain unclear, most retinal disease variants map onto the DEAD domain (**Fig. 2b-c**), consistent with a role for the DEAD domain in ATP hydrolysis and RNA binding and remodeling^19^. Using predicted DDX41 structure, the mutant, p.I396T (M1), would disrupt a central hydrophobic pocket that stabilizes the DEAD domain and would likely compromise stability of the protein (**Fig. 2c-d**). Consistent with this prediction, overexpression of DDX41 p.I396T in HEK293F cells yields in 8-10-fold less protein than wild-type, and the purified protein fails to bind a heparin column, a proxy for RNA binding (**Fig. 2e-f**), indicating disrupted folding and impaired RNA interaction. By contrast, the p.R339C variant is expressed at levels comparable to wild-type but displays a distinct profile on heparin chromatography (**Fig. 2e-f**), consistent with modified RNA-binding properties and modestly reduced stability. Supporting this, increased amounts of the Hsp70 chaperone co-purify with p.R339C (**Fig. 2g**). While previous work reported strong DDX41 binding to DNA and DNA:RNA hybrids^20^, here we observed moderately strong binding to RNAs longer than ∼20 nucleotides. Electrophoretic Mobility Shift Assay (EMSA) experiments show that while one or two copies of wild-type DDX41 bind RNA in the absence of ATP, the p.R339C mutant decreases DDX41 binding to RNA as distinct single- or double-bound complexes (**Fig. 2h-j**). Instead, an accumulation of higher-order multimeric RNA-bound complexes is observed for the p.R339C protein, consistent with formation of aggregates or potentially partially misfolded DDX41 multimers that can still bind RNA (**Fig. 2h**). Altogether, our biochemical studies indicate that retinal diseases variants disrupt the ability of DDX41 to interact with RNA, either through misfolding or through formation of non-productive higher-order assemblies that may lead to dysfunctional aggregation on RNA (**Fig. 2j**).

**Fig. 2.**
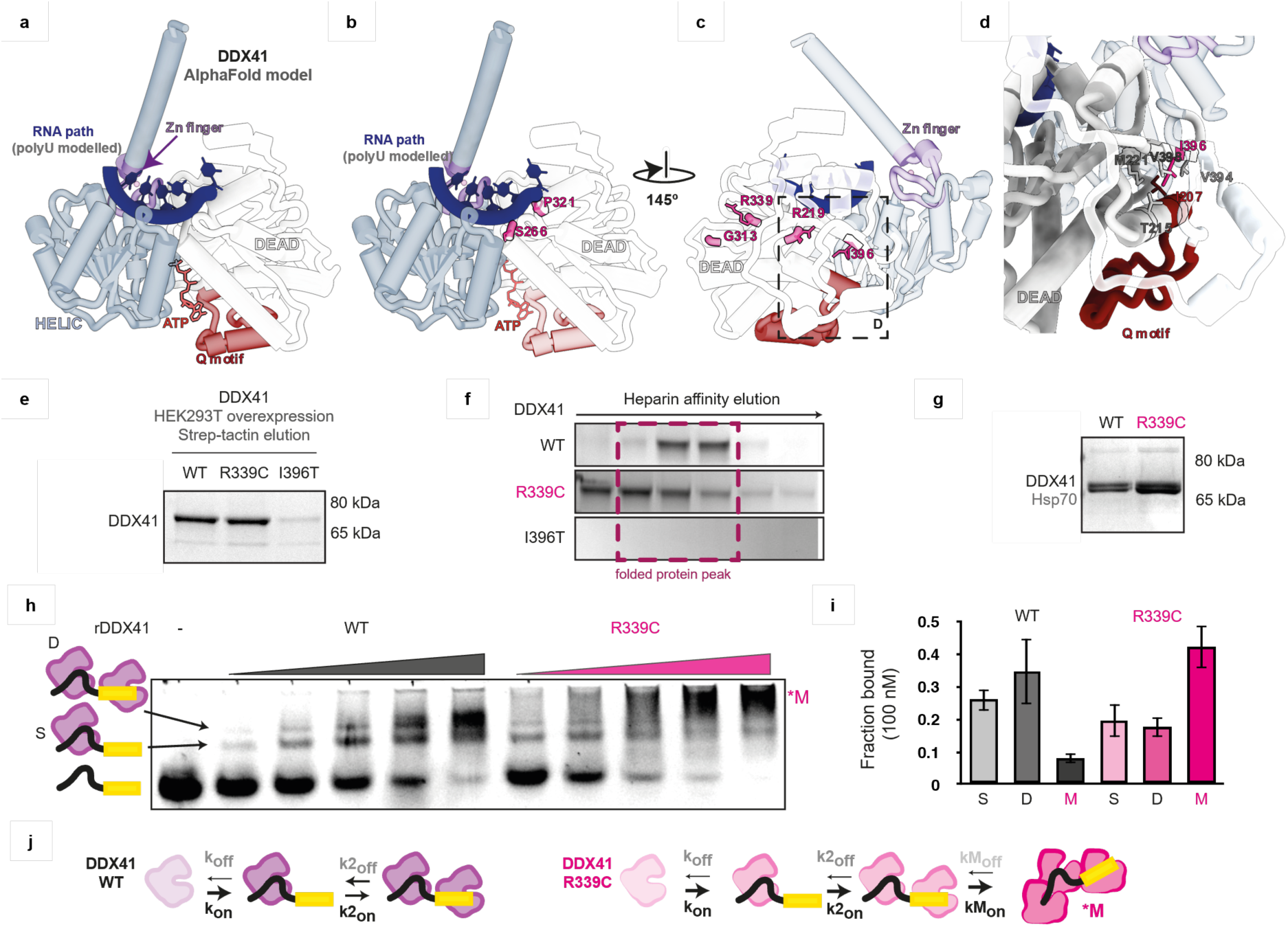
Structural modeling and biochemical analysis of DDX41 variants. (a) Predicted 3D structure of human DDX41 bound to poly-uridine RNA, modeled with AlphaFold3^61^ and adjusted based on Dbp5-RNA structure^62^; N-terminal region truncated for clarity. (b-d) Locations of *DDX41* variants on the DEAD domain, highlighting residues predicted to affect RNA binding (S266, P321) and the role of I396 in stabilizing the hydrophobic pocket and Q-motif. Models were visualized in ChimeraX^63^. (e-g) Expression and purification of DDX41 variants from HEK293F cells. Proteins were visualized by (e) Coomassie staining, (f) after Heparin/StrepTactin affinity purification, by silver staining and (g) immunoblot of purified proteins for EMSA using an anti-DDX41 antibody shows co-purification of Hsp70, with increased association observed for the p.R339C variant, supporting partial misfolding of this mutant. Hsp70 represents a contaminant co-purifying with DDX41 rather than a specific target of the antibody. (h-j) EMSA analysis of DDX41 binding to RNA and structural/kinetic model. (h) Representative EMSA blot showing single (S) and double (D) and higher order (M) DDX41-RNA complexes that form sequentially. Schematic on the left depicts the molecular species producing the observed bands: free Cy5-labeled RNA (RNA shown as a black ribbon, Cy5 as a yellow box), RNA bound by a single (S) DDX41 (purple) and by two (D) DDX41 molecules. (i) Quantification of bound fractions at 100 nM protein; error bars represent standard deviation from at least two independent purifications (n = 4). The R339C variant shows a marked decrease in single-bound complexes and a concomitant increase in double-bound species, consistent with disrupted RNA binding and partial misfolding. (j) Kinetic model for DDX41-RNA binding, with a schematic depicting single (S), double (D), and higher-order (M) assemblies. Kon/Koff and K2on/K2off indicate association/dissociation rates for S and D complexes, respectively, while KMon/KMoff correspond to higher-order assemblies. Higher-order complexes may represent either aggregates of multiple DDX41 monomers or multimeric assemblies that bind RNA less specifically.

### The Homozygous c.1187T>C (M1) Variant Triggers Proteasomal Degradation of DDX41 and R-loop Dysregulation

Fibroblasts from individuals P1 and P2 carrying the homozygous M1 variant exhibited markedly reduced DDX41 protein levels in patient cells compared with controls in total lysates as well as cytoplasmic and nuclear fractions (**Fig. 3a-b**; **Supplementary Fig.2a-d**). Treatment with the proteasome inhibitor MG132 partially restored DDX41 levels, implicating proteasomal degradation, supported by accumulation of ubiquitinated proteins following MG132 exposure (**Fig. 3c-d**; **Supplementary Fig. 2e**). By contrast, lysosomal inhibition by Bafilomycin A1 (BafA1) had no effect, ruling out lysosomal degradation (**Supplementary Fig. 2f-g**). Immunofluorescence analysis showed that DDX41 colocalizes with nuclear speckle marker SC35, also known as Serine/Arginine-Rich Splicing Factor 2 (SRSF2), consistent with a role in RNA processing; however, nuclear DDX41 was significantly diminished in patient fibroblasts compared with controls, in agreement with Western blot analyses (**Fig. 3e-f**). Consistent with impaired DDX41 function, patient fibroblasts displayed a marked increase in nuclear RNA-DNA hybrids compared to controls, as detected by S9.6 immunofluorescence, indicating elevated R-loop levels (**Supplementary Fig.2h-i**). These findings are in line with previous studies implicating DDX41 in the maintenance of R-loop homeostasis^7^.

**Fig. 3.**
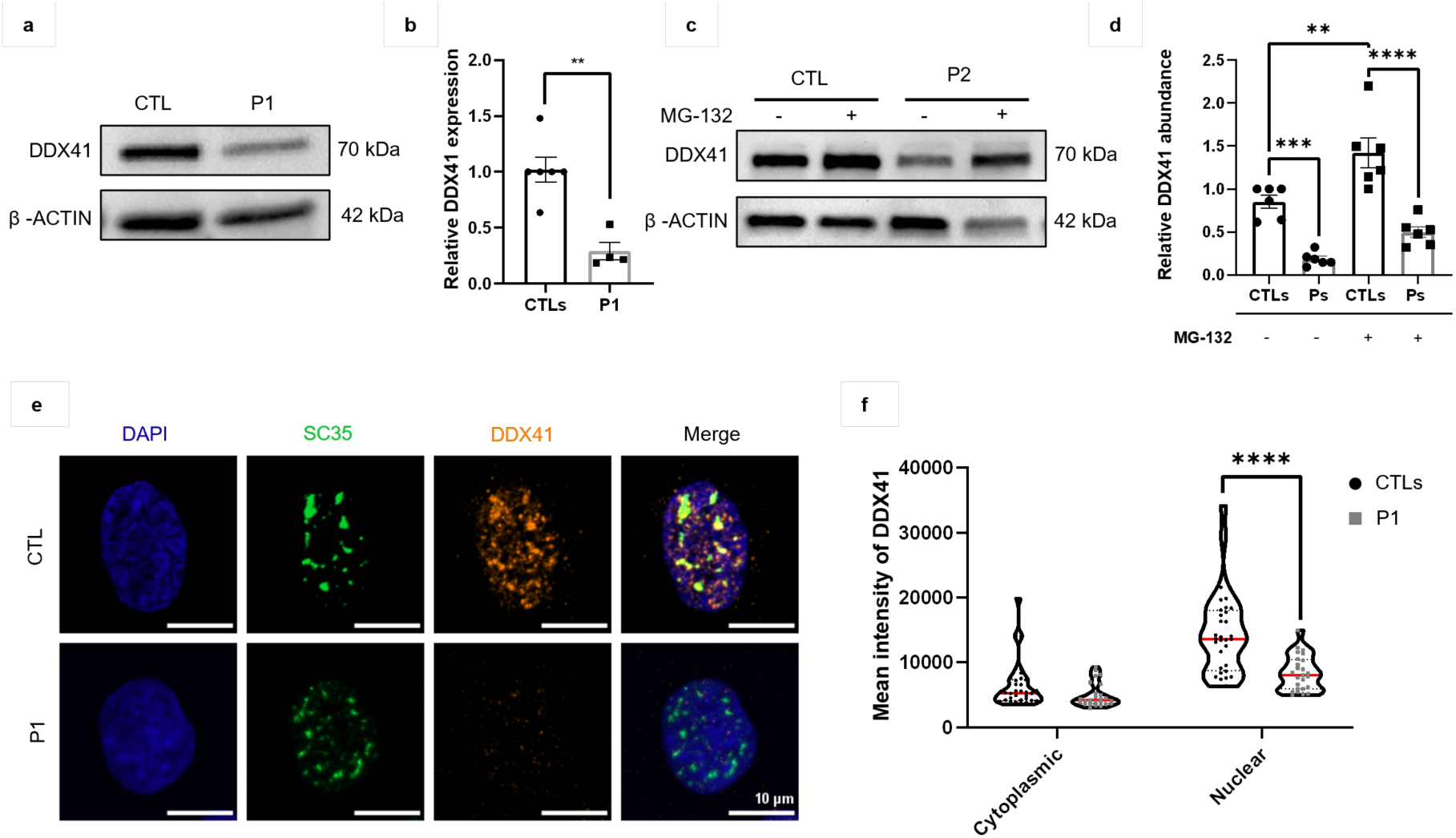
DDX41 expression, stability, and subcellular localization in control and patient fibroblasts. (a, b) DDX41 protein levels in total cell lysates from control (CTL) and patient (P1) fibroblasts. (a) Representative Western blot; β-actin was used as a loading control. Molecular weight markers are shown on the right. (b) Quantification of DDX41 levels normalized to β-actin. Data are mean ± SEM from three independent experiments. **, p = 0.0013 (unpaired two-tailed Student t-test). (c, d) DDX41 expression in control (CTL) and patients (Ps, P1-P2) fibroblasts after treatment with 10 μM MG-132 for 18 h. (c) Representative Western blot; β-actin is the loading control. (d) Quantification of DDX41 levels in treated and untreated cells, normalized to β-actin. Data represent mean ± SEM from three independent experiments. **, p = 0.0039, ***, p = 0.0008, ****, p < 0.0001 (two-way ANOVA with Tukey post hoc test). (e, f) Immunofluorescence analysis of DDX41 (orange) and the nuclear speckles marker SC35 (green) (n=3). (e) Representative images of control and patient fibroblasts; merged images show predominantly nuclear DDX41 with partial colocalization with SC35. Scale bars, 10 µm. (f) Quantification of DDX41 fluorescence intensity in cytoplasmic and nuclear compartments. Data represent mean ± SEM from three independent experiments; ****, p < 0.0001 (two-way ANOVA with Tukey post hoc test).

### Loss of DDX41 Function Results in Immune-related Transcriptional Reprogramming and Splicing Defects, with snRNA Dysregulation Trends

RNA-seq analysis of fibroblasts from patient P1 and four controls revealed largely unchanged *DDX41* mRNA levels in patient cells (**Fig. 4a**). Differential expression analysis (False Discovery Rate (FDR) < 0.05; Fold Change (FC), |log₂FC| ≥ 1.5) identified 174 differentially expressed genes (DEGs), including 43 downregulated and 131 upregulated genes (**Fig. 4b-c**). Gene Ontology (GO) enrichment analysis revealed an overrepresentation of genes related to extracellular matrix, negative regulation of cell proliferation, vasculature development, response to growth factors, and cytokine activity. Among the most upregulated genes, we identified those involved in immune response and inflammation (e.g. *CD36, CD4, SERPINB2*^21–23)^, various aspects of cancer biology, including tumor progression, metastasis, angiogenesis, and cell survival pathways (e.g. *ADGRF5, MYH11, FOXS1, BEX1*^24–27)^ and transcriptional regulation (e.g. *ZNF423, SOBP*^28,29^), consistent with previous reported functions of DDX41^16,30^.

**Fig. 4.**
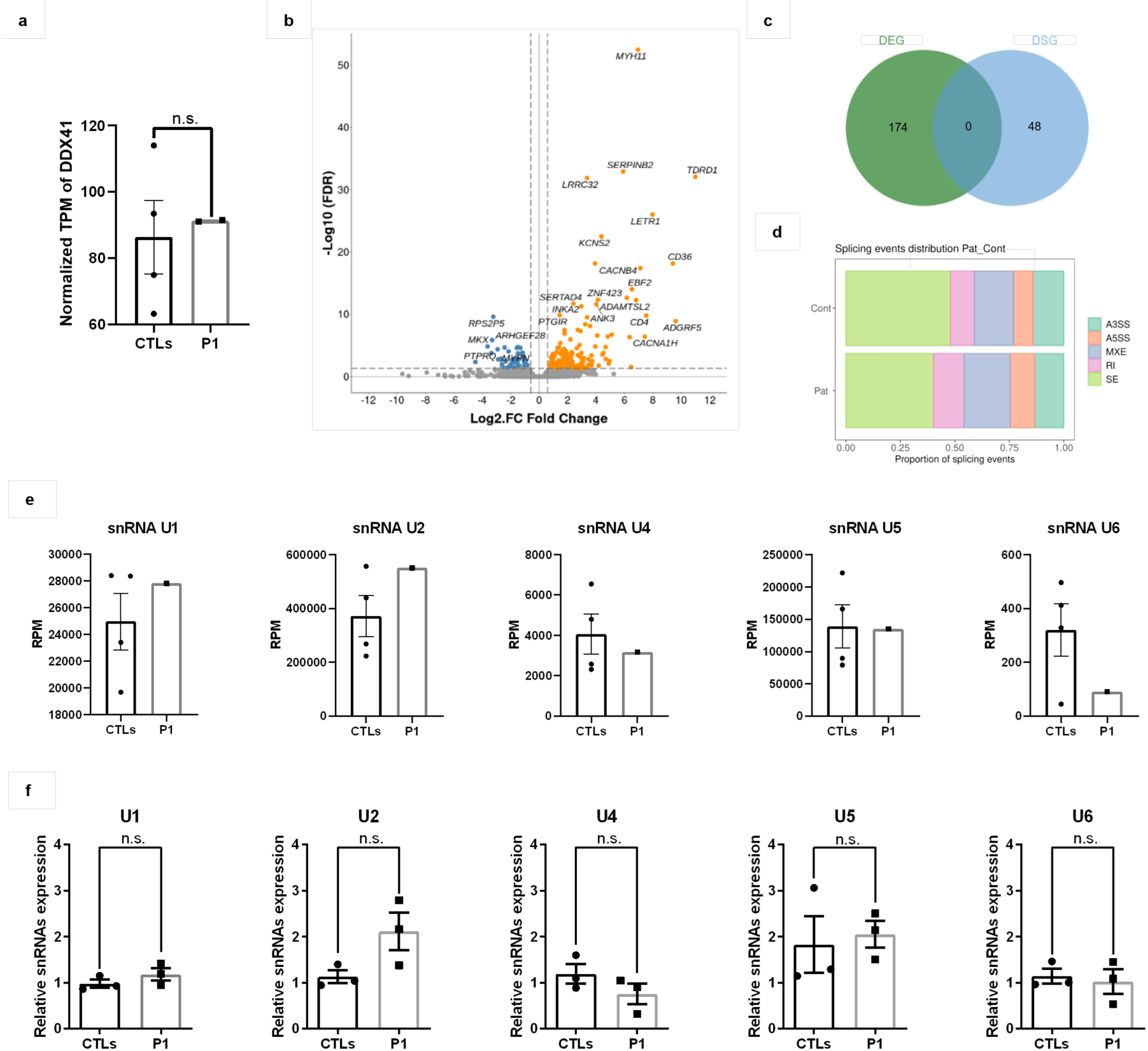
Whole-transcriptome analysis in control and patient fibroblasts. (a) DDX41 transcript levels were comparable between the affected individual (P1) and healthy controls (CTLs), as determined by RNA-seq. Bars represent mean ± SEM; n.s., not significant (unpaired two-tailed Student t-test). (b) Volcano plot of differentially expressed genes (DEGs) between patient and control fibroblasts. The x-axis shows log₂ fold change (FC) in gene expression; the y-axis represents statistical significance as -log₁₀ adjusted false discovery rate (FDR). Genes with |log₂FC| ≥ 1.5, TPM ≥ 5, and FDR < 0.05 were considered significant. Significantly upregulated genes are shown in orange, downregulated genes in blue. (c) Venn diagrams depicting the overlap between DEGs and differentially spliced genes (DSGs) identified in patient versus control fibroblasts. (d) Distribution of splicing event types based on rMATS analysis. Skipped exons (SE) were the most frequent event, followed by alternative 5′ splice sites (A5SS), alternative 3′ splice sites (A3SS), retained introns (RI), and mutually exclusive exons (MXE). (e) RNA-seq quantification of spliceosomal small nuclear RNAs (snRNAs) U1, U2, U4, U5, and U6 in fibroblasts from the affected individual (P1) and controls (CTLs), expressed as reads per million (RPM). Bars represent mean ± SEM. (f) RT-qPCR validation of U1, U2, U4, U5, and U6 snRNA levels in fibroblasts from four controls and the affected individual. Bars represent mean ± SEM from three independent experiments; n.s., not significant (unpaired Welch t-test was used to assess statistical differences).

Analysis of differential isoform usage identified 63 splicing alterations across 48 genes (FDR < 0.05), predominantly involving reduced skipped exons (SEs) and increased mutually exclusive exons (MXEs) (**Fig. 4c-d**). Notably, Differentially Spliced Genes (DSGs) did not overlap with DEGs, indicating splicing changes independent of transcriptional modulation. Several DSGs participated in immune-related pathway and GO analysis highlighted enrichment for DEAD/H-box RNA helicase binding and negative regulation of the Fas signaling pathway, emphasizing alternative splicing as a distinct regulatory layer in cellular and immune functions.

RNA-seq analysis of gel-purified small nuclear RNAs (snRNAs) revealed comparable abundances of *U1*, *U2*, *U4*, *U5*, and *U6* snRNAs in patient and control fibroblasts (**Fig. 4e**), which was validated by semiquantitative RT-qPCR (**Fig. 4f**). A modest trend toward increased *U2* snRNA was observed, potentially reflecting a compensatory response or subtle dysregulation in RNA processing pathways linked to *DDX41* dysfunction.

### Mice Expressing *Ddx41* M1-equivalent *(Ddx41^I396T/I396T^)* Exhibit Early ERG Deficit and Progressive Photoreceptor Degeneration

CRISPR/Cas9 knock-in mouse model harboring the human M1 variant (*Ddx41^I396T/I396T^*) were viable and exhibited no overt morphological, developmental, or behavioral abnormalities (**Fig. 5a**). Given the retinal defects observed in human patients (**Supplementary Table 2**), we assessed retinal function and anatomy longitudinally in *Ddx41^I396T/I396T^* and *Ddx41^WT/WT^* mice. ERG analysis revealed significant reductions in scotopic (rod-mediated) and photopic (cone-mediated) a- and B-wave amplitudes in *Ddx41^I396T/I396T^* mice from 1 month onward, indicating impaired photoreceptor function and compromised inner retinal signaling (**Fig. 5b**). Histological analysis from 1 to 12 months revealed gradual retinal degeneration, with a marked reduction in outer nuclear layer (ONL) nuclei in *Ddx41^I396T/I396T^* mice at 12 months, indicating progressive age-dependent retinal loss (**Fig. 5c-e**).

**Fig. 5.**
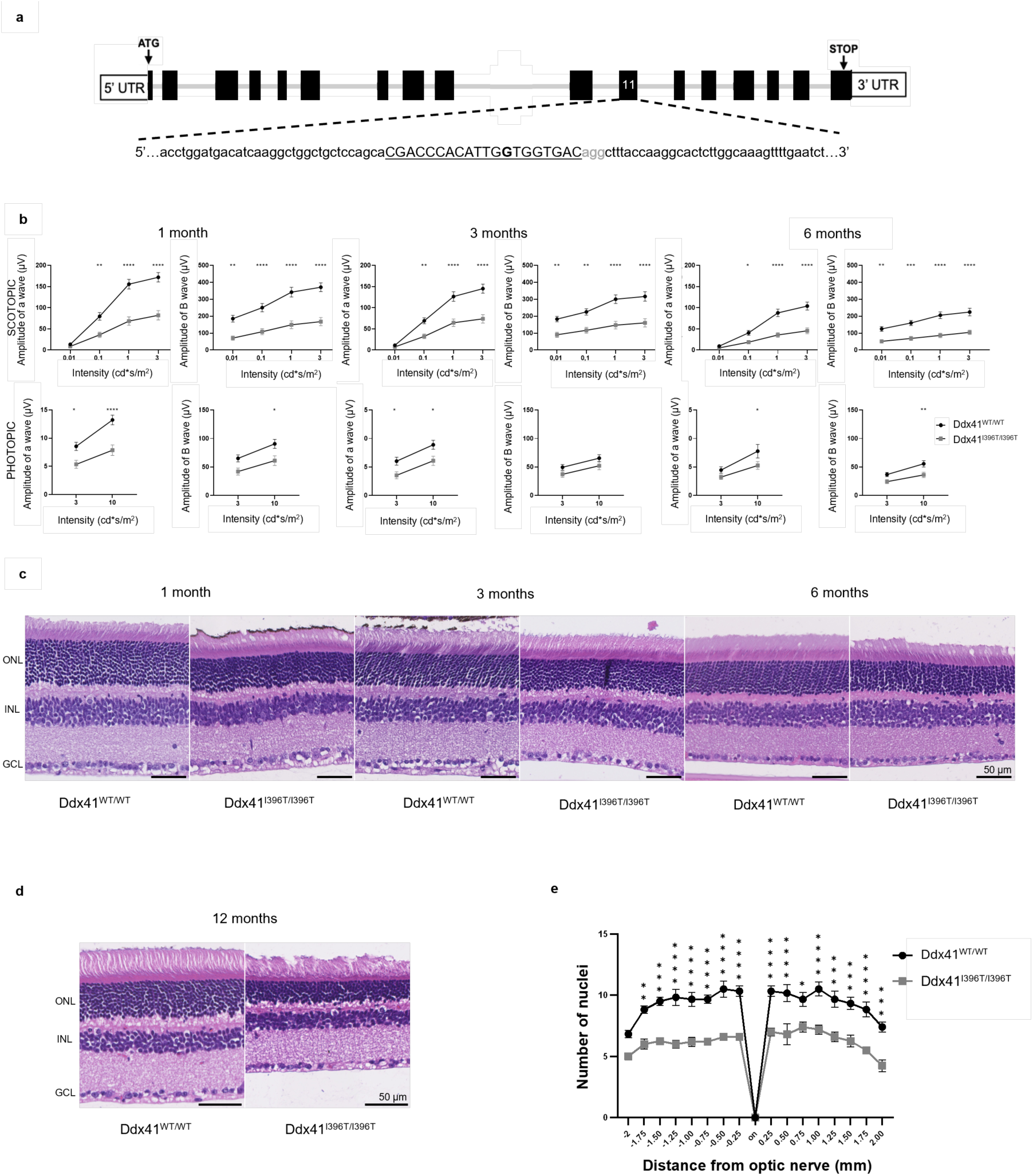
Functional and structural characterization of the *Ddx41^I396T/I396T^* knock-in mouse model. (a) Schematic of the CRISPR-Cas9 genome editing strategy used to generate the *Ddx41*^I396T/I396T^ mouse. Coding exons are shown as solid black bars, non-coding exons as open bars. The sgRNA sequence (black underline) targets exon 11, spanning the isoleucine codon at position 396 (I396). The protospacer adjacent motif (PAM) is shown in grey, and the introduced nucleotide substitution is indicated in bold. (b) Electroretinography (ERG) analysis of cone- and rod-mediated responses in wild-type (WT) and *Ddx41*^I396T/I396T^ mice at postnatal months 1, 3, and 6. Dot plots show a-wave and B-wave amplitudes. Statistical significance between age-matched genotypes was assessed by two-way ANOVA followed by Sidak post hoc test. Bars represent mean ± SEM from n = 10 mice per genotype per time point. Significance: *: p ≤ 0.05; **: p ≤ 0.01; ***: p ≤ 0.001; ****: p ≤ 0.0001. (c) Representative retinal histology images of WT and *Ddx41*^I396T/I396T^ mice at postnatal months 1, 3, and 6 (40× magnification). Retinal layers appear intact in both genotypes across all time points. ONL: Outer Nuclear Layer; INL: Inner Nuclear Layer; GCL: Ganglion Cell Layer. (d) Retinal histology at postnatal month 12 showing thinning of the ONL and reduced photoreceptor nuclei in *Ddx41*^I396T/I396T^ mice compared with WT. (e) Quantification of ONL nuclei counts per 250 µm retinal length at postnatal month 12. Each dot represents one biological replicate (n = 4-6 retinas per genotype). Statistical analysis was performed using two-way ANOVA with Sidak correction for multiple comparisons. Bars represent mean ± SEM. Significance: *: p ≤ 0.05; **: p ≤ 0.01; ***: p ≤ 0.001; ****: p ≤ 0.0001.

### Alterations in the Retinal Transcriptome Precede Structural Degeneration and Dysregulate Visual Perception and Müller Glia Pathways

RNA-seq analysis of 1-month-old *Ddx41^I396T/I396T^* retinas, a stage at which the retina shows no overt degeneration, revealed a total of 256 DEGs compared with their wild-type littermates (p-value <0.01 and |log₂FC| ≥ 1.2) (**Fig. 6a**). GO analysis highlighted enrichment for genes implicated in visual perception, including retinoid cycle components, eye development, and branching, suggesting that *Ddx41* influences retinal development and structural organization. Notably, genes from Müller cell repertoire as *Cd44*, *Rdh10*, *Rgr*, *Socs3* and *Pdgfr3* were dysregulated (**Fig. 6b**). Differential isoform analysis showed 17 splicing events in 8 genes (*Ppp4c, Serf2, Rnf123, Laptm4a, Mrps17, Saxo2, Gas5* and *Gm57315*), predominantly alternative 5’ splice sites (A5SS), indicating a context- and tissue-specific role for *Ddx41* in splicing (**Supplementary Table 3)**.

**Fig. 6.**
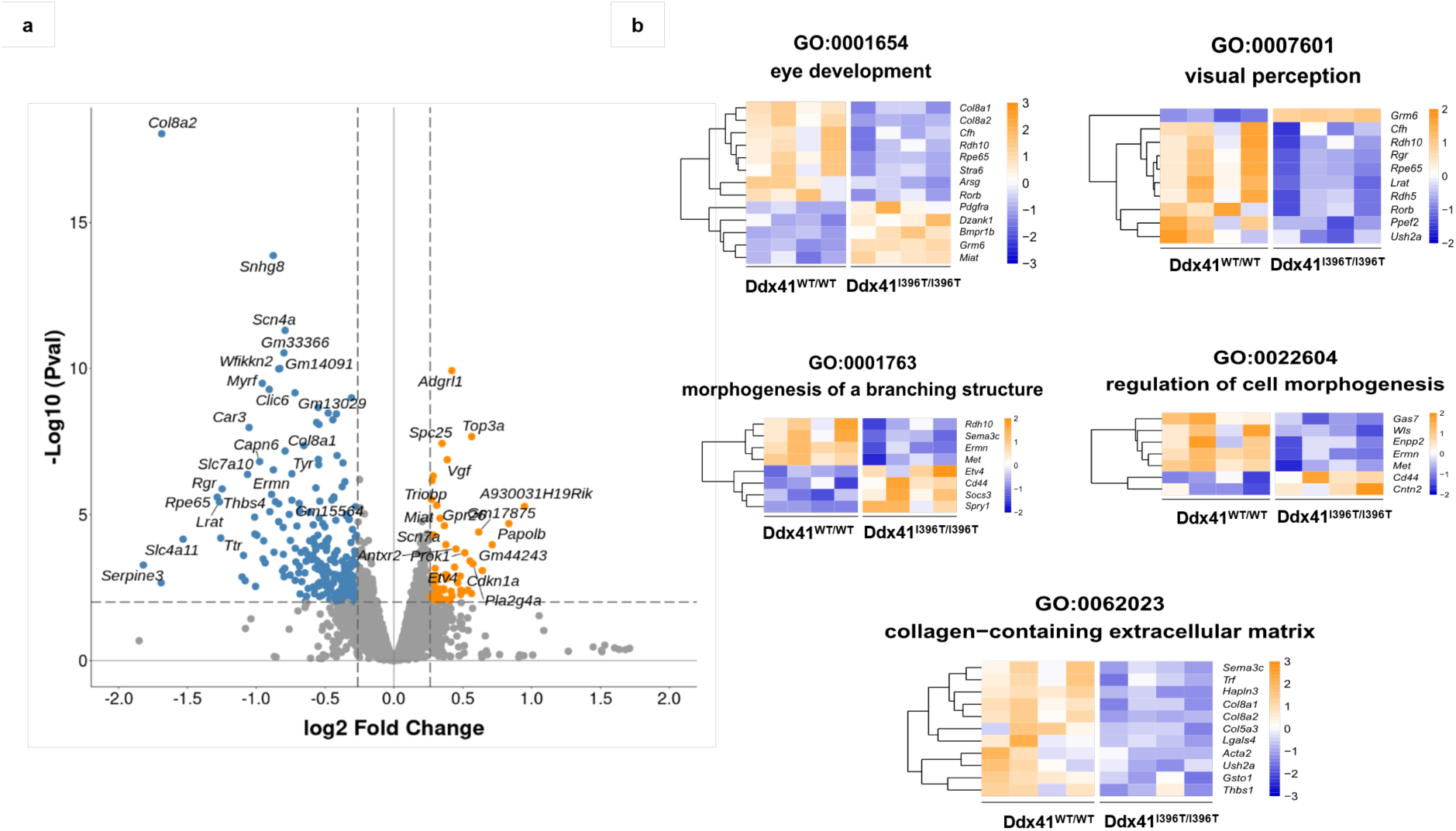
Transcriptomic profiling of retinas from 1 month-old wild-type and *Ddx41^I396T/I396T^* mice. (a) Volcano plot of differentially expressed genes (DEGs) between wildtype (WT) and *Ddx41^I396T/I396T^* mutant retinas (n = 4 per genotype) at postnatal month 1. The x-axis shows log₂ fold change (FC) in gene expression, and the y-axis shows statistical significance as -log₁₀ FDR-adjusted p-values. Genes with |log₂FC| ≥ 1.2, TPM ≥ 5, and P < 0.01 were considered significant. Upregulated genes are shown in orange, downregulated genes in blue. (b) Heatmaps generated using Metascape showing enrichment of Gene Ontology (GO) biological pathways among DEGs. Each row represents a gene set, and color intensity reflects relative expression levels (blue: low expression; orange: high expression).

### Proteomic Analysis of *Ddx41^I396T/I396T^*Retina Supports Structural Dysfunction

Comparative mass spectrometry of total lysates of retinas from 1-month-old *Ddx41^I396T/I396T^* mice and wild-type littermates identified 103 differentially expressed proteins (DEPs; FDR < 0.05 and |log₂FC| ≥ 1.5), including 61 upregulated and 42 downregulated proteins (**Fig. 7a**).

**Fig. 7.**
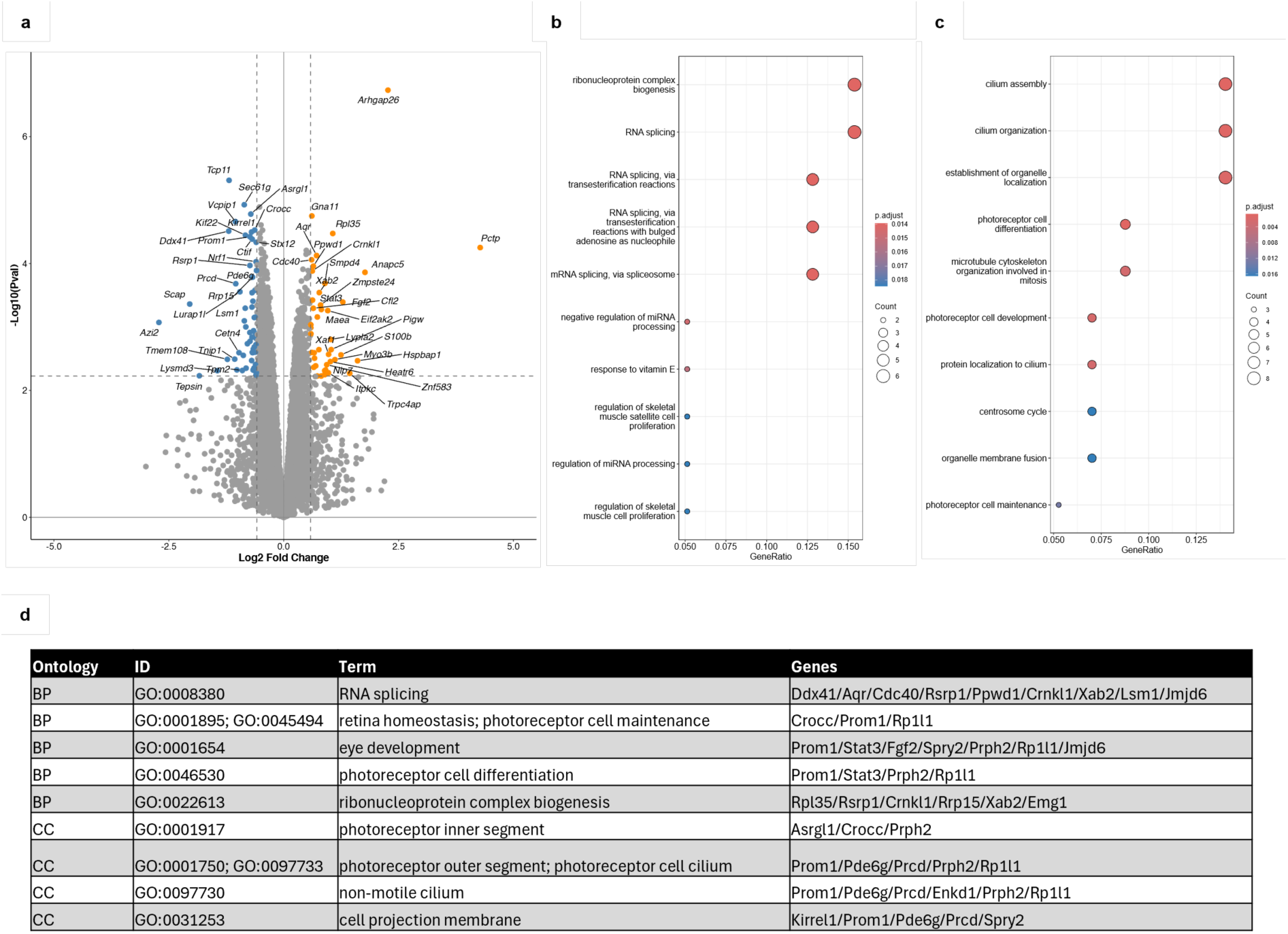
Proteomic analysis of from 1 month-old wild-type and *Ddx41^I396T/I396T^* mice. (a) Volcano plot of proteins significantly upregulated (orange) or downregulated (blue) *Ddx41^I396T/I396T^* mutant *versus* wild-type (WT) retinas (n = 3 per genotype) at postnatal month 1. Differentially expressed proteins were defined as |log₂FC| ≥ 1.5 and FDR ≤ 0.05. (b) Dot plots showing Gene Ontology (GO) biological pathway enrichment among upregulated proteins. Pathways related to ribonucleoprotein complex biogenesis and RNA splicing are prominently represented. (c) Dot plots showing GO biological pathway enrichment among downregulated proteins. Pathways related to cilium assembly, organization, and photoreceptor cell differentiation are prominently represented. (d) Table displaying GO enrichment analysis of the 50 most significantly altered proteins (FDR < 0.05). BP: Biological Process; CC: Cellular Component.

The downregulated proteins were enriched for pathways essential for photoreceptor structure and function, including outer segment formation, cilium assembly and photoreceptor cell differentiation, whereas upregulated proteins were associated with RNA splicing and ribonucleoprotein complex biogenesis pathways (**Fig. 7b-c**). Gene Ontology enrichment analysis of the 50 most significantly altered proteins highlighted retinal homeostasis, non-motile cilium assembly, photoreceptor outer/inner segment organization, and RNA processing/splicing, reflecting the broad impact of DDX41 on retinal cell biology (**Fig. 7d**). Notably, several adhesion proteins including CRB1, CRB2, and other junctional components were downregulated in mutant retinas, suggesting the disruption of the integrity of adherent junctions and extracellular matrix contacts at the outer limiting membrane, where Müller glial endfeet interface with photoreceptors. This finding aligns with transcriptomic evidence of Müller cell impairment and supports the hypothesis that mutant DDX41 compromises retinal structural integrity partly by affecting Müller cell adhesion and function.

DDX41 was among the downregulated proteins, a finding confirmed by Western blot analysis showing reduced DDX41 levels in *Ddx41^I396T/I396T^*retinas compared with wild-type controls (**Fig. 8a-b**). Immunolabeling further revealed nuclear DDX41 throughout the retina in both genotypes at 8 months, with highest expression in the outer nuclear layer (ONL), inner nuclear layer (INL), and ganglion cell layer (GCL) (**Fig. 8c**). A similar nuclear distribution was observed in human retinal sections, with strongest expression in the ONL and INL (**Supplementary Fig.3**), highlighting conservation across species.

**Fig. 8.**
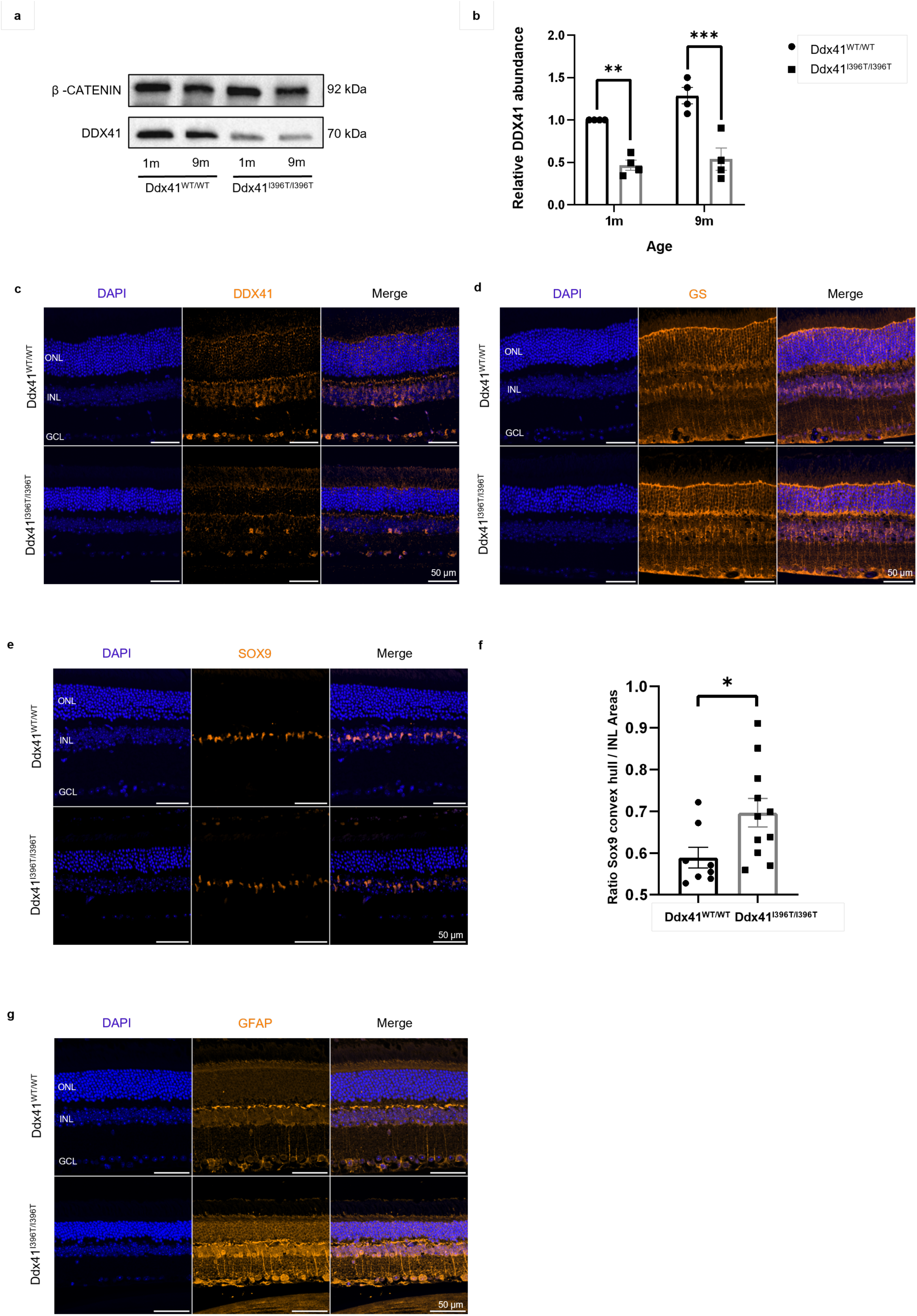
Immunolabeling of DDX41 and Müller glia markers in the retina of wild-type and *_Ddx41_I396T/I396T* mice. (a, b) Western blot analysis and quantification of DDX41 expression in retinal protein extracts from wildtype (WT) and *Ddx41^I396T/I396T^* mice (n = 4 per genotype) at 1 and 9 months of age. (a) Representative Western blot; β-catenin was used as a loading control, and molecular weight markers are indicated on the right. (b) Quantification of DDX41 levels normalized to β-catenin, showing reduced expression in *Ddx41^I396T/I396T^*mice compared with WT at both time points. Data represent mean ± SEM, **, p = 0.0042, ***, p = 0.0002 (two-way ANOVA with Tukey post hoc test). (c-f) Immunolabeling and quantitative analysis of DDX41 and Müller glia organization in retinas from 8-month-old WT and *Ddx41^I396T/I396T^* mice. (c-e) Immunofluorescence staining showing (c) DDX41 (orange), (d) glutamine synthetase (GS; cytoplasmic Müller glia marker), and (e) SOX9 (nuclear Müller glia-specific transcription factor). Scale bars, 50 µm. (f) Quantification of Müller glia nuclear spatial dispersion within the inner nuclear layer (INL) based on SOX9 staining between WT and *Ddx41*^I396T/I396T^. The ratio between the convex hull area encompassing SOX9-positive nuclei and the total INL area was calculated using QuPath (version 0.5.1)^64^. This dimensionless index reflects glial spatial organization, with higher values indicating broader nuclear dispersion. Bars represent mean ± SEM (n = 3 per genotype). *, p = 0.0307 (unpaired two-tailed t-tests). (g) GFAP immunostaining of retinal sections from 12-month-old WT and *Ddx41^I396T/I396T^*mice, indicating gliosis. Scale bar, 50 μm.

### Müller Glial Cell Spatial Organization and Architecture Altered in *Ddx41^I396T/I396T^* Retina

Immunostaining of 8-month-old *Ddx41^I396T/I396T^*and wild-type retinas revealed altered distribution and organization of Müller glial cells. Glutamine synthetase (GS), a cytoplasmic marker of Müller glia involved in neurotransmitter recycling and metabolic support, suggested a tendency toward Müller cell disorganization, together with increased GS labeling (**Fig. 8d**). SOX9, a Müller glia-specific nuclear transcription factor marking Müller glial nuclei, suggested subtle alterations in Müller cell nuclei spatial distribution and organization. This observation was supported by automated spatial analysis showing a shift in clustering, consistent with a disruption of Müller glial organization associated with the *Ddx41* variant (**Fig. 8e-f**). GFAP immunostaining at 12 months further confirmed reactive gliosis in Müller cells from *Ddx41^I396T/I396T^* retinas, with increased staining intensity (**Fig. 8g**) Together, these findings associate DDX41-dependent proteomic alterations and reduced protein abundance with structural changes in Müller glia, supporting a model in which DDX41 dysfunction may compromise retinal integrity through impaired Müller cell adhesion and organization.

### *Ddx41^I396T/I396T^* Mice Display Preserved Hematopoiesis at Steady State and Aging-Related Defects After Transplantation

Complete blood counts (CBC) in 11-week-old *Ddx41^I396T/I396T^* mice and wild-type littermates revealed no significant differences in the white blood cell (WBC), red blood cell (RBC), platelet (PLT) counts (**Supplementary Fig. 4a-c**), indicating normal peripheral blood composition at this age. Bone marrow analysis revealed no change in the total number of cells (**Supplementary Fig. 4d**). Looking at progenitor populations, including long-term (LT) and short-term (ST) hematopoietic stem cells, multipotent progenitors (MPP), common myeloid progenitors (CMP), granulocyte-macrophage progenitors (GMP), and megakaryocyte-erythroid progenitors (MEP), by FACS also revealed no differences between genotypes (Supplementary **Fig. 4e-f**), suggesting preserved progenitor distribution and function.

Donor bone marrow transplanted *Ddx41^I396T/I396T^* mice (BMT) were longitudinally monitored from 4 to 10 months. Peripheral blood cell counts remained stable (**Supplementary Fig. 4g-j**). Bone marrow analysis revealed an age-associated trend toward increased hematopoietic stem cells (HSCs) with concomitant decreases in multipotent progenitor 2 cells (MPP2) and megakaryocyte-erythroid progenitor (MEP) populations, consistent with normal hematopoietic aging^31,32^ (**Supplementary Fig. 4k-l**). Together, these results indicate that *Ddx41^I396T/I396T^* mice maintain normal hematopoiesis under both homeostatic and transplantation conditions.

## DISCUSSION

Recessive germline variants in *DDX41* define a syndrome characterized by hereditary retinal dystrophy, variable neurodevelopmental involvement, and craniofacial and skeletal anomalies. At least one previously described individual^15^ also exhibits retinal phenotype, indicating that ocular involvement may have been under-recognized in prior reports. Affected individuals display a broad spectrum of neurological features, ranging from mild balance and coordination difficulties to cerebellar ataxia, psychomotor delay and, in some cases, febrile seizures. Skeletal manifestations include short stature, brachydactyly, microcephaly, and other craniofacial anomalies^16^. Collectively, these findings delineate a coherent multi-system phenotype and expand the spectrum of DDX41-associated disease beyond its established role in hematologic malignancies^11,33,34^.

This study identified eight germline variants, six of which located within the DEAD domain of the protein. Three are frameshift mutations (M3, M4 and M5). The M3 variant (in coiled coil domain), is observed in trans with a missense variant, while the M4 (in DEAD domain), and M5 frameshift variants are also in trans, a configuration expected to result in a complete loss of functional protein. Notably, no homozygous truncating variants are reported in gnomAD v4.1.0 (probability of Loss-of-function Intolerance; pLI = 0), consistent with the non-viability of complete loss of function, as supported by the embryonic lethality of germline *Ddx41* knockout mice^35^. We suspected that alternative transcripts may escape Nonsense-Mediated Decay (NMD) and produce sufficient functional protein for viability despite these truncating events, although this cannot be confirmed without patient-derived material.

Mechanistically, DDX41 displays a conserved nuclear DEAD-box helicase-like architecture with critical roles in maintaining genomic stability, including resolution of R-loops and modulation of RNA-protein interactions^5,7,36,37^. Loss of DDX41 results in R-loop accumulation and activation of DNA damage responses, processes that are particularly deleterious in post-mitotic retinal cells, likely contributing to photoreceptor dysfunction and degeneration^38,39^. DDX41 has been previously implicated in pre-mRNA splicing and found associated with spliceosome complexes^40,41^. Our transcriptomic data further indicated a stable small nuclear RNA (snRNA) landscape, though we observed potential shifts in the processing or abundance in *snRNA U2* that align with DDX41 known processing roles^12,42^. In our RNA-seq data, only a limited number of differential splicing events and relatively few misspliced genes were detected in mutant compared with wild-type mouse retinas. Nonetheless, DDX41 is a critical regulator of retinal splicing. Mutations in core spliceosomal components can selectively disrupt specific steps of the splicing process without collapsing the entire machinery, as demonstrated for yeast Brr2 (ortholog of human SNRP200) ^43^ and human PRP8^44^. These observations suggest that DDX41 variants may selectively affect discrete splicing events and transcript subsets, sufficient to drive severe retinal pathology.

Retinal dysfunction is further modulated by Müller glial cells, as suggested by transcriptomic alterations in the *Ddx41^I396T/I396T^*mouse model phenotypes, absent from prior lab models^45^. These glial cells are essential for retinal homeostasis and support of photoreceptors, and their dysfunction may amplify retinal pathology^46–48^. Together, these findings establish DDX41 as a key regulator of retinal cellular homeostasis, linking its loss to tissue-specific vulnerability.

Interestingly, retinal and neurological phenotypes were sometimes precipitated or exacerbated by systemic infections in patients carrying the p.I396T variant. This observation is consistent with the role of microglia in mediating inflammatory responses, where systemic infection can trigger retinal inflammation and contribute to neurodevelopmental or degenerative symptoms^49–53^. Such gene-environment interactions illustrate how external stressors can modify the penetrance and severity of DDX41-related phenotypes, particularly in tissues with heightened sensitivity to genomic stress. Recent work by Sharma et al. (2024)^16^ provides further, albeit indirect, support for this hypothesis by showing that germline *DDX41* variants distinct from p.I396T, impair activation of interferon-stimulated genes via the STING-type I interferon pathway in patient-derived fibroblasts. While this points to a potential role for altered innate immune signaling in DDX41-associated disease, comparison with our fibroblast RNA-seq data revealed limited transcriptomic convergence, with only 58 shared deregulated genes among the 968 reported by Sharma et al.^16^, and no deregulation of their proposed key candidate POSTN. These differences suggest that immune pathway perturbations may be variant-specific and/or highly context dependent, emphasizing the need for future studies assessing DDX41 function under inflammatory conditions in disease-relevant neural and retinal cell types.

The genotype-phenotype relationship of *DDX41* variants is further nuanced by tissue context. Retinal disease appears to be driven solely by germline variants, whereas hematologic malignancies typically follow a classic two-hit model, requiring both a germline variant and a subsequent somatic event in hematopoietic cells to fully disrupt tumor suppressor function^54^. Germline *DDX41* pathogenic variants are associated with a late-onset predisposition to myeloid neoplasms such as MDS and AML, with most diagnoses occurring in the sixth to seventh decade of life and lifetime penetrance that can be substantial compared with the general population (∼50% by late adulthood)^54^. Large cohorts of DDX41-related myeloid malignancies show a pronounced male predominance (often ∼3:1 male:female), despite similar germline carrier frequencies between sexes, suggesting the involvement of sex-specific modifiers of leukemogenesis^17^. An important mechanistic consideration relates to the well-characterized DDX41 p.R525H variant, which frequently arises as a somatic “second hit” in heterozygous carriers and is strongly associated with progression to myeloid malignancy^3,10,11^. Functional studies indicate that p.R525H impairs DDX41 helicase activity and fails to rescue loss-of-function in hematopoietic cells, consistent with a hypomorphic effect rather than a gain-of-function^55^. Among the older relatives of the patients reported here, only one individual from Family 7, whose DDX41 status could not be determined due to unavailable DNA, is known to have developed a hematologic malignancy. It remains possible that additional cases exist among older relatives, but this cannot be determined at present, underscoring the need to collect further data from older individuals. Importantly, early-stage hematologic malignancies may be clinically silent, with peripheral blood counts remaining within normal ranges despite ongoing disease development; detection may therefore require bone marrow evaluation and careful monitoring for cytopenia. Continued longitudinal follow-up-including periodic hematologic assessment-will be essential to detect potential late-onset manifestations in all *DDX41* germline variants carriers^56^.

The individuals described here carry biallelic loss-of-function variants (5 males, 6 females; median age 27 ± 13.5 years) and have no overt hematologic malignancy up to 45 years of age, with the exception of one case who developed BPDCN in the first decade (Family 9)^15^. Notably, analysis of leukemic blasts from this individual revealed no additional DDX41 mutations and the older genoidentical sibling of this individual has no overt symptoms of BPDCN^15^. The occurrence of hematologic malignancy in one sibling but not the other suggests that biallelic loss-of-function variants alone are not sufficient to cause leukemic transformation, and that additional genetic, epigenetic, or environmental factors may contribute to malignancy. Whether other individuals carrying germline biallelic DDX41 mutations have an increased risk of hematologic malignancy remains to be determined.

Overall, our study establishes DDX41 as a critical factor for retinal homeostasis and neurodevelopment, while extending its well-characterized role in hematopoiesis to the retina. The identification of a germline-driven retinal dystrophy expands the phenotypic spectrum of DDX41-associated disease and emphasizes the importance of considering this gene in genetic diagnosis of inherited retinal dystrophies^57^. Mechanistically, these findings highlight the role of RNA helicase function, genomic instability, and glial-mediated support in tissue-specific pathology, providing a framework to understand how DDX41 variants produce divergent outcomes in different organs. Future studies will be critical to dissect the molecular pathways linking DDX41 dysfunction to retinal degeneration, neurodevelopmental impairment, and hematologic risk, potentially uncovering novel therapeutic avenues for these multi-system disorders.

Collectively, these findings place DDX41 within the broader landscape of RNA helicases critical for retinal function. Other DEAD/H-box helicases, including DDX3x and DHX38, have been shown to regulate retinal development and function, with variants causing early-onset retinal dystrophies^58–60^. This emerging pattern underscores the importance of RNA helicase-mediated RNA processing and genome maintenance in photoreceptor survival and highlights how defects within this family result in tissue-specific degeneration.

## Data Availability

All relevant data, detailed protocols, antibody RRIDs, and analysis code are available upon request.

## ACKNOWLEDGEMENTS

This work was supported by grants from Retina France, Fondation Visio and Inflam’oeil to J.M.R. and I.P.. SL is supported by the Medical Research Council (Clinician Scientist Fellowship, grant reference UKRI440), and the National Institute for Health and Care Research (NIHR) Manchester Biomedical Research Centre (BRC) (NIHR203308). This work was supported by grants from the National Eye Institute [EY022356, EY018571]] and Retinal Research Foundation to R.C.. M.J.H.H. discloses support from Knights Templar Eye foundation (grant 2024-09). C.A. was supported by Instituto de Salud Carlos III (ISCIII) of the Ministerio de Ciencia e Innovación and Unión Europea - European Regional Development Fund (FEDER) (grant nos. PI22/00321 and IMP/00009), Centro de Investigación Biomédica en Red Enfermedades Raras (CIBERER, grant no. 06/07/0036), IIS-FJD BioBank (grant no. PT23/00114), the Organización Nacional de Ciegos Españoles (ONCE), the European Regional Development Fund (FEDER) and the University Chair UAM-IIS-FJD of Genomic Medicine. This work was performed by using the data contained in the ‘Programa Infraestructura de Medicina de Precisión asociada a la Ciencia y la Tecnología en Medicina Genómica (IMPaCT-GENóMICA)’, coordinated by the CIBERER and founded by ISCIII.

J.M.M. received three grants from Instituto de Salud Carlos III (ISCIII), “PI22/00213”, “AC21_2/00022” and FORT23/00021 co-funded by the European Union, and the grant CIPROM/2023/26 from the Generalitat Valenciana. GGG acknowledges two grants from Instituto de Salud Carlos III (ISCIII), “CP22/00028” and “PI22/01371”, co-funded by the European Union. GGG has also a grant funded by the European Union in the HORIZON programme HORIZON-HLTH-2023-TOOL-05-04 (BETTER, 101136262). PBM received a grant from Ministerio de Universidades “FPU20/04736”. P.Y. was supported by the National Institutes of Health (Bethesda, MD) P30 EY010572 core grant, the Malcolm M. Marquis, MD Endowed Fund for Innovation, and an unrestricted grant from Research to Prevent Blindness (New York, NY) to Casey Eye Institute, Oregon Health & Science University.

The authors thank Necker Bioimage Analysis (Nicolas Goudin), Cell Imaging (Meriem Garfa Traoré), Histology (Sofian Ameur, Damien Conrozier, Mayeul Thomas and Sophie Berissi), Genomic and Bioinformatic platforms (Nicolas Cagnard) for valuable discussions and help. Thanks go to Edwin L.Van Dick for performing small nuclear RNAs RNASeq. Special thanks to the European Retinal disease consortium.

The authors would like to acknowledge all patients and their families for participating in this study.

## AUTHOR CONTRIBUTIONS

Z.M performed in vitro and in vivo experiments, analyzed the data, and wrote the paper. A.Z, K.K, T.M, V.M, D.L, A.A, C.C, L.G, H.H.M.J, A.R.M., L.Y, X.B, C.Z performed experiments. P.D. and L.F-T. performed creation of animal models. G.A, I.A, C.A, P.B-M, B.B, A.B-C, H.D, L.F-C, G.G-G, S.L, E.M, J.M.M, A.R.M, M.P.M-G, M.P.R, S.T, S.T-S, I.M, L.K, K.A.W.W, A.R.W, M.E.P, P.Y collected clinical data. J.E.R. analyzed data from RNASeq. R.S-L. analyzed data from snRNASeq. C.I.G. and V.J. performed mass spectrometry. H.D collected skin biopsies from patients. I.P. analyzed exome data from Family 1 and Family 2. K.K, M.Q analyzed data from Family 3, 4 and 5. M.W analyzed exome data from Family 6. O.P analyzed genome data from Family 7. S.G, R.C, F.R-L, S.M.F, M.S supervised the research. C.R supervised the research and participated actively to the writing. J-M.R. and I.P. supervised the research, designed the experiments, and wrote the paper. All authors discussed the results and participated in manuscript preparation and editing.

## MATERIAL & METHODS

### Families

Thirteen affected individuals (five males and eight females) from nine unrelated families diagnosed with visual symptoms consistent with retinal dysfunction were included in this study.

Family 1 comprised two affected siblings (P1-P2) born to consanguineous parents. Family 2 included one affected individual (P3), with no reported consanguinity. Family 3 was a consanguineous family with two affected siblings (P4-P5). Family 4 had one affected individual (P6), with no reported consanguinity. Family 5 included one affected individual (P7) without reported consanguinity. Family 6 consisted of two affected siblings (P8 and P9), with documented consanguinity. Family 7 involved one affected individual (P10), with no known consanguinity. Family 8 included one affected individual (P11), with varying degrees of consanguinity. Finally, Family 9 comprised two affected siblings, with no reported consanguinity; these cases were previously described by Diness et al. (2018)^15^.

All participants or their legal guardians provided written informed consent in accordance with the Declaration of Helsinki. Ethics approval was obtained from the institutional review boards of all participating centers.

### *DDX41* Variants Identification

Variants in *DDX41* were identified after excluding pathogenic variants in known inherited retinal dystrophy (IRD) genes through analysis of a curated IRD gene panel^57^. Subsequently, whole-exome sequencing (WES) was performed for Families 1, 2, and patient P8 (Family 6); whole-genome sequencing (WGS) was carried out for patients P4 and P5 (Family 3), P6 (Family 4), P7 (Family 5), P10 (Family 7), and P11 (Family 8); trio genome sequencing was performed within the *Plan France Médecine Génomique* (PFMG2025) framework for Family 5; and Sanger sequencing was used for patient P9.

Genomic DNA libraries for patients P1, P2, and P3 were prepared using the SureSelectXT Library Prep Kit (Agilent) following DNA shearing with a Covaris S2 Ultrasonicator. Target regions of interest (ROIs) were captured with the SureSelect All Exon V5 kit (Agilent) and sequenced on an Illumina HiSeq2500 HT platform. Data analysis was performed through a custom bioinformatics pipeline (POLYWEB) developed by the Imagine Institute Bioinformatics Core, Paris Descartes University.

P6 underwent WGS after KAPA Hyper Prep PCR-free library preparation and sequencing on a NovaSeq 6000 system (Illumina). Raw data were processed using the DNASeq bioinformatics pipeline available at https://github.com/TBLabFJD/PARROT-FJD, applying BWA alignment, GATK best practices, and Ensembl VEP annotation. P8 was analyzed using WES with the Twist Human Core Exome Library on NovaSeq X (150PE, 6 Gb raw data), following the procedure described in PMID 36909829. P9 was analyzed by Sanger sequencing.

Variant pathogenicity predictions were obtained using PolyPhen-2, SIFT, and MutationTaster algorithms, while allele frequencies were referenced against the gnomAD population database. Segregation analysis in the parents of affected individuals was performed by Sanger sequencing, confirming biparental inheritance of *DDX41* variants in most cases. Variant annotation was based on the GRCh38 human genome assembly and GenBank transcript NM_016222.4.

### Fibroblast Tissue Collection and Cell culture

Primary dermal fibroblasts were established from skin biopsies of two affected individuals, P1 and P2 (Family 1), as well as from four unrelated healthy control subjects (C1-C4, CTLs). Cultures were maintained in Opti-MEM GlutaMAX I medium (Life Technologies) supplemented with 10% fetal bovine serum (FBS; Life Technologies), 50 U/mL penicillin, and 50 μg/mL streptomycin (Life Technologies).

Human embryonic kidney (HEK293F) cells were cultured in DMEM/F-12 medium (Life Technologies) supplemented with 10% FBS and 1% penicillin/streptomycin. All cell lines were maintained in a humidified incubator at 37 °C with 5% CO₂.

### Expression and Purification of Recombinant DDX41 for Biochemical Assays

Available on request.

### Heparin Chromatography assay of DDX41 variants

Available on request.

### Electrophoretic Mobility Shift Assay (EMSA)

Available on request.

### Generation of Ddx41^I396T/I396T^ Mouse Model and Ethics

A mouse model carrying the *DDX41* p.I396T missense variant: C57BL/6J - *Ddx41^em(I396T)^* which will be name *Ddx41^I396T/I396T^*in this study, orthologous to the human variant, was generated via CRISPR-Cas9-mediated genome editing at the laboratory of Animal experimentation and Transgenesis facility (LEAT) of the Institut Imagine. A single guide RNA (sgRNA) targeting the exon 11 regions of the mouse *Ddx41* gene near the I396 codon was designed using CRISPOR software and synthesized by Integrated DNA Technologies (IDT). A single-stranded oligodeoxynucleotide (ssODN) repair template encoding the desired c.1187T>C nucleotide change (resulting in I396T amino acid substitution) along with silent mutations to prevent re-cutting was synthesized (Figure 5a). Newborn mice were genotyped using genomic DNA extracted from tail biopsies, amplified by PCR, and confirmed by Sanger sequencing with specific primers (Eurofins, Forward 5’-CCCCCAAAGGGGTTGAGACC-3’, and Reverse 5’-GGGAGCAAGGCAGCTTTAGCC-3’). The generated animals were backcrossed to a C57BL/6J background before use in experiments.

For breeding, *Ddx41^I396T/WT^* animals were intercrossed to obtain *Ddx41^I396T/I396T^* homozygotes and wild-type littermates. All animal procedures were approved by the French Ministry of Research (APAFIS#47246) and complied with institutional animal care and ethical guidelines of the Animal Care and Use Committee of Paris Descartes University and the LEAT Facility, Institut Imagine.

### Electroretinographic Analysis of Mouse Model

Retinal function in *Ddx41^I396T/I396T^*mice and wild-type littermates aged 1 to 6 months was assessed by electroretinography (ERG) using the Celeris system (Diagnosys LLC), following previously described protocols^66^.

### Western Blot Analysis

Protein extracts from cell cultures were prepared as previously described^67^ using samples from two independent controls and two patient-derived fibroblast lines. Membranes were incubated overnight at 4 °C with a polyclonal rabbit anti DDX41 antibody (1:2000; HPA017911, Merck), followed by incubation with an HRP conjugated goat anti rabbit IgG secondary antibody (1:10,000; Invitrogen). Immunoblots were developed and analyzed as reported in Barny et al. (2018)^67^. DDX41 signal intensity was normalized to β actin (1:10,000; ab8226, Abcam).

For tissue analysis, 1-month-old and 9-month-old *Ddx41^I396T/I396T^*and wild-type mice (n = 3 per genotype) were euthanized by cervical dislocation, and eyes were enucleated. Retinas were lysed in RIPA buffer supplemented with protease and phosphatase inhibitor cocktail (PIC; Thermo Fisher Scientific). Western blotting was performed as described above, except that DDX41 abundance across samples was normalized to β-catenin, using a polyclonal goat anti β-catenin primary antibody (1:500; AF1329, R&D Systems) and an HRP conjugated donkey anti goat IgG secondary antibody (1:10,000; Invitrogen). Quantitative data represent the mean ± SEM from three independent protein extractions.

### MG132 Treatment and Western Blot Analysis

Primary dermal fibroblasts from two independent controls and two patient-derived fibroblast lines were treated with MG132 (474790, Merck) at 10 µM for 18 h to inhibit the proteasome prepared in DMSO and diluted into culture medium, with control cells receiving equivalent DMSO vehicle. Treatment conditions were based on prior optimization and relevant literature^16^.

Western blot analysis was performed as described in the previous section. Treatment efficacy was verified mouse monoclonal anti-ubiquitin (1:1000, 3936S, Cell Signaling) for proteasome inhibition. DDX41 was detected with primary and HRP-conjugated secondary antibodies followed by chemiluminescent detection.

### Immunofluorescence, Immunohistochemistry and Imaging Analyses

For immunofluorescence analysis, primary dermal fibroblasts from two independent controls and two patients were seeded at a density of 2 × 10⁵ cells per well on glass coverslips in 12 well plates. Cells were fixed with 100% methanol and blocked with 3% bovine serum albumin (BSA) and 0.1% Triton X 100 in phosphate buffered saline (PBS). DDX41 was immunostained overnight at 4 °C using the following primary antibodies: polyclonal rabbit anti DDX41 (1:200; HPA017911, Merck), monoclonal mouse anti SC 35 (1:2000; S4045, Sigma Aldrich), and monoclonal mouse anti DNA/RNA hybrid (clone S9.6, 1:200; MABE1095, Merck). Secondary labeling was performed for 1 h at room temperature using Alexa 555 conjugated donkey anti rabbit IgG (1:1000; Thermo Fisher Scientific) and Alexa 488 conjugated donkey anti mouse IgG (1:1000; Thermo Fisher Scientific).

For immunohistochemistry, deparaffinized eye sections (n = 3 per genotype) were subjected to antigen retrieval in 10 mM trisodium citrate buffer (pH 6.0) containing 0.05% Tween 20 for 30 min at 95 °C. Sections were blocked for 1 h in PBS containing 5% BSA and incubated overnight at 4 °C in a humidified chamber with rabbit anti DDX41 (1:50; HPA017911, Merck) diluted in blocking solution. Slides were washed three times in PBS and incubated for 1 h at room temperature with Alexa 555 conjugated donkey anti rabbit IgG (1:200; Thermo Fisher Scientific).

Additional murine eye sections were immunostained using the following primary antibodies: polyclonal rabbit anti SOX9 (1:1000; ab185966, Abcam), polyclonal rabbit anti Glutamine Synthetase (GS) (1:200; ab168020, Abcam), and polyclonal rabbit anti Glial Fibrillary Acidic Protein (GFAP) (1:100; Z0334, Dako). Detection was carried out with Alexa 555 conjugated donkey anti rabbit IgG (1:200; Thermo Fisher Scientific).

Nuclei were counterstained with DAPI (Invitrogen), and sections were mounted in Fluoromount medium (Sigma) under glass coverslips. Images were acquired using a Zeiss Spinning Disk microscope equipped with 20× air, 40×/1.3 oil, or 63×/1.4 oil objectives.

Quantitative analysis of DDX41, S9.6, and SOX9 staining was performed using QuPath (version 0.5.1)^64^ and a machine-learning-based classification to distinguish true signal. The mean intensities of nuclear and cytoplasmic DDX41, nuclear S9.6, and nuclear SOX9 staining, as well as SOX9 signal density, were measured. Final image panels were prepared using FigureJ on Fiji (version 1.54)^68,69^.

### Proteomics Sample Preparation and Mass Spectrometry

Retina tissues from both wild-type (n =3) and *Ddx41^I396T/I396T^* (n = 3) mice at 1 month of age were lysed for 1 hour in 100 μL RIPA buffer supplemented with protease and phosphatase inhibitors (PIC; Thermo Fisher Scientific). Samples were sonicated to ensure complete protein solubilization, and protein concentrations were determined using the Bradford assay (Thermo Fisher Scientific).

Sample processing, digestion, and peptide clean-up were performed according to the protocol described by Zanetti et al. (2024)^45^. Quantitative mass spectrometry analyses were conducted as previously reported, with detailed acquisition parameters and bioinformatics workflows described in Zanetti et al. (2024)^45^.

The raw proteomics data have been deposited in the ProteomeXchange Consortium under the identifier PXD071486.

### RNA Isolation, Sequencing, Differential Expression and Pathway Analysis

Total RNA was isolated from duplicate fibroblast cultures of one patient and four independent controls, as well as retinal tissues (n = 4 per genotype), using the RNeasy Mini Kit (Qiagen), followed by on column DNase I treatment with the RNase Free DNase Set (Qiagen). RNA concentration, purity, and integrity were assessed with an Agilent 2100 Bioanalyzer (Agilent Technologies).

For RNA-Seq analysis, retinas from wild-type (n =4) and *Ddx41^I396T/I396T^*(n = 4) mice at 1 month of age were used processed. RNA Seq library preparation and sequencing were performed as previously described^45^. Read quantification was carried out using *featureCounts*^70^ to generate gene level count tables.

Differential expression analyses were conducted with the *edgeR* package^71^ to perform normalization, statistical testing, and calculation of TPM (transcripts per million) values. Differentially expressed genes (DEGs) were defined by a fold change >|1.2|, P < 0.01, and minimum expression level ≥ 5 TPM.

Analyses of alternative splicing, pathway enrichment, and gene annotation were performed following the pipelines described by Zanetti et al. (2024)^45^. Raw RNA Seq data have been deposited in the BioStudies repository under accession identifiers S-BSST2316 (for fibroblast RNA Seq) and S-BSST2318 (for retina RNA Seq).

### Quantitative Analysis of U1, U2, U4, U5, U6 snRNA Abundance in Patient and Control Fibroblasts

snRNA from P1 and four independent controls fibroblasts were lysed using TRIzol and extracted with the Direct-zol RNA MiniPrep Plus (Qiagen and Zymo Research, respectively). RT-qPCR analysis and transcriptomic analysis of small nuclear RNAs (snRNAs) was performed as previously described in ref^45^. The raw data have been deposited in BioStudies ArrayExpress with the identifier E-MTAB-16362.

### Statistical Analysis

Statistical parameters including the exact sample size (n), post hoc tests, and statistical significance are reported in every figure and figure legend. Data were estimated to be statistically significant when p ≤ 0.05 by Student t-test, one or two-way ANOVA. Statistical analyses were performed using GraphPad PRISM (v8) (GraphPad Software Inc., La Jolla, CA, USA). All values are shown as mean ± SEM. Source data are provided as a Source data excel file.

## REFERENCES

1. Ali, M. A. M. The DEAD-box protein family of RNA helicases: sentinels for a myriad of cellular functions with emerging roles in tumorigenesis. Int. J. Clin. Oncol. 26, 795–825 (2021).

2. Cargill, M., Venkataraman, R. & Lee, S. DEAD-Box RNA Helicases and Genome Stability. Genes 12, 1471 (2021).

3. Polprasert, C. et al. Inherited and Somatic Defects in DDX41 in Myeloid Neoplasms. Cancer Cell 27, 658–670 (2015).

4. Chlon, T. M. et al. Germline DDX41 mutations cause ineffective hematopoiesis and myelodysplasia. Cell Stem Cell 28, 1966–1981.e6 (2021).

5. Kadono, M. et al. Biological implications of somatic DDX41 p.R525H mutation in acute myeloid leukemia. Exp. Hematol. 44, 745–754.e4 (2016).

6. Saruul Tungalag et al. Ribosome profiling analysis reveals the roles of DDX41 in translational regulation. Int. J. Hematol. 117, 876–888 (2023).

7. Mosler, T. et al. R-loop proximity proteomics identifies a role of DDX41 in transcription-associated genomic instability. Nat. Commun. 12, 7314 (2021).

8. Shinriki, S. et al. DDX41 coordinates RNA splicing and transcriptional elongation to prevent DNA replication stress in hematopoietic cells. Leukemia 36, 2605–2620 (2022).

9. Omura, H. et al. Structural and Functional Analysis of DDX41: a bispecific immune receptor for DNA and cyclic dinucleotide. Sci. Rep. 6, 34756 (2016).

10. Sébert, M. et al. Germline DDX41 mutations define a significant entity within adult MDS/AML patients. Blood 134, 1441–1444 (2019).

11. Duployez, N. et al. Prognostic impact of DDX41 germline mutations in intensively treated acute myeloid leukemia patients: an ALFA-FILO study. Blood 140, 756–768 (2022).

12. Makishima, H., Bowman, T. V. & Godley, L. A. DDX41-associated susceptibility to myeloid neoplasms. Blood 141, 1544–1552 (2023).

13. Bannon, S. A. et al. Next-Generation Sequencing of DDX41 in Myeloid Neoplasms Leads to Increased Detection of Germline Alterations. Front. Oncol. 10, 582213 (2021).

14. Lewinsohn, M. et al. Novel germ line DDX41 mutations define families with a lower age of MDS/AML onset and lymphoid malignancies. Blood 127, 1017–1023 (2016).

15. Diness, B. R. et al. Putative new childhood leukemia cancer predisposition syndrome caused by germline bi-allelic missense mutations in *DDX41*. Genes. Chromosomes Cancer 57, 670–674 (2018).

16. Sharma, P. et al. Biallelic germline DDX41 variants in a patient with bone dysplasia, ichthyosis, and dysmorphic features. Hum. Genet. 143, 1445–1457 (2024).

17. Kim, K., Ong, F. & Sasaki, K. Current Understanding of DDX41 Mutations in Myeloid Neoplasms. Cancers 15, 344 (2023).

18. Schütz, P. et al. Comparative Structural Analysis of Human DEAD-Box RNA Helicases. PLoS ONE 5, e12791 (2010).

19. Mallam, A. L., Del Campo, M., Gilman, B., Sidote, D. J. & Lambowitz, A. M. Structural basis for RNA-duplex recognition and unwinding by the DEAD-box helicase Mss116p. Nature 490, 121–125 (2012).

20. Singh, R. S. et al. DDX41 is required for cGAS-STING activation against DNA virus infection. Cell Rep. 39, 110856 (2022).

21. Luckheeram, R. V., Zhou, R., Verma, A. D. & Xia, B. CD4^+^T cells: differentiation and functions. Clin. Dev. Immunol. 2012, 925135 (2012).

22. Schroder, W. A., Major, L. & Suhrbier, A. The Role of SerpinB2 in Immunity. Crit. Rev. Immunol. 31, 15–30 (2011).

23. Silverstein, R. L. & Febbraio, M. CD36, a scavenger receptor involved in immunity, metabolism, angiogenesis, and behavior. Sci. Signal. 2, re3 (2009).

24. Alhopuro, P. et al. Somatic mutation analysis of MYH11 in breast and prostate cancer. BMC Cancer 8, 263 (2008).

25. Jacenik, D., Hikisz, P., Beswick, E. J. & Fichna, J. The clinical relevance of the adhesion G protein-coupled receptor F5 for human diseases and cancers. Biochim. Biophys. Acta Mol. Basis Dis. 1869, 166683 (2023).

26. Lee, S. et al. BEX1 and BEX4 Induce GBM Progression through Regulation of Actin Polymerization and Activation of YAP/TAZ Signaling. Int. J. Mol. Sci. 22, 9845 (2021).

27. Liu, Y., Tu, M. & Wang, L. Pan-Cancer Analysis Predicts FOXS1 as a Key Target in Prognosis and Tumor Immunotherapy. Int. J. Gen. Med. 15, 2171–2185 (2022).

28. Harder, L., Puller, A.-C. & Horstmann, M. A. ZNF423: Transcriptional modulation in development and cancer. Mol. Cell. Oncol. 1, e969655 (2014).

29. Tavares, A. L. P., Jourdeuil, K., Neilson, K. M., Majumdar, H. D. & Moody, S. A. Sobp modulates the transcriptional activation of Six1 target genes and is required during craniofacial development. Dev. Camb. Engl. 148, dev199684 (2021).

30. Winstone, L., Jung, Y. & Wu, Y. DDX41: exploring the roles of a versatile helicase. Biochem. Soc. Trans. 52, 395–405 (2024).

31. Amoah, A. et al. Aging of human hematopoietic stem cells is linked to changes in Cdc42 activity. Haematologica 107, 393–402 (2021).

32. Young, K. et al. Progressive alterations in multipotent hematopoietic progenitors underlie lymphoid cell loss in aging. J. Exp. Med. 213, 2259–2267 (2016).

33. Badar, T. et al. Clinical and molecular correlates of somatic and germline *DDX41* variants in patients and families with myeloid neoplasms. Haematologica 108, 3033–3043 (2023).

34. Badar, T. & Chlon, T. Germline and Somatic Defects in DDX41 and its Impact on Myeloid Neoplasms. Curr. Hematol. Malig. Rep. 17, 113–120 (2022).

35. Ma, J., Mahmud, N., Bosland, M. C. & Ross, S. R. DDX41 is needed for pre- and postnatal hematopoietic stem cell differentiation in mice. Stem Cell Rep. 17, 879–893 (2022).

36. Jiang, Y. et al. Structural and functional analyses of human DDX41 DEAD domain. Protein Cell 8, 72–76 (2017).

37. Weinreb, J. T. et al. Excessive R-loops trigger an inflammatory cascade leading to increased HSPC production. Dev. Cell 56, 627–640.e5 (2021).

38. Jauregui-Lozano, J. et al. Proper control of R-loop homeostasis is required for maintenance of gene expression and neuronal function during aging. Aging Cell 21, e13554 (2022).

39. Sun, K. et al. The splicing factor DHX38 enables retinal development through safeguarding genome integrity. iScience 26, 108103 (2023).

40. Dybkov, O. et al. Regulation of 3’ splice site selection after step 1 of splicing by spliceosomal C* proteins. Sci. Adv. 9, eadf1785 (2023).

41. Osterhoudt, K., Bagno, O., Katzman, S. & Zahler, A. M. Spliceosomal helicases DDX41/SACY-1 and PRP22/MOG-5 both contribute to proofreading against proximal 3′ splice site usage. RNA 30, 404–417 (2024).

42. Hershberger, C. E., Daniels, N. J. & Padgett, R. A. Spliceosomal factor mutations and mis-splicing in MDS. Best Pract. Res. Clin. Haematol. 33, 101199 (2020).

43. Zhao, C. et al. Autosomal-Dominant Retinitis Pigmentosa Caused by a Mutation in SNRNP200, a Gene Required for Unwinding of U4/U6 snRNAs. Am. J. Hum. Genet. 85, 617–627 (2009).

44. Zimmann, F. et al. Retinitis pigmentosa-linked mutations impair the snRNA unwinding activity of SNRNP200 and reduce pre-mRNA binding of PRPF8. Cell. Mol. Life Sci. 82, 103 (2025).

45. Zanetti, A. et al. GPATCH11 variants cause mis-splicing and early-onset retinal dystrophy with neurological impairment. Nat. Commun. 15, 10096 (2024).

46. Li, J., Patil, R. V. & Verkman, A. S. Mildly abnormal retinal function in transgenic mice without Müller cell aquaporin-4 water channels. Invest. Ophthalmol. Vis. Sci. 43, 573–579 (2002).

47. Nag, T. C. Müller cell vulnerability in aging human retina: Implications on photoreceptor cell survival. Exp. Eye Res. 235, 109645 (2023).

48. Bachleda, A. R., Pevny, L. H. & Weiss, E. R. Sox2-Deficient Müller Glia Disrupt the Structural and Functional Maturation of the Mammalian Retina. Invest. Ophthalmol. Vis. Sci. 57, 1488–1499 (2016).

49. Blackburn, K. M. & Wang, C. Post-infectious neurological disorders. Ther. Adv. Neurol. Disord. 13, 1756286420952901 (2020).

50. Kelly, R., Reinert, L. S. & Paludan, S. R. Sequelae of viral CNS infections including outcomes, mechanisms, and knowledge gaps. Npj Viruses 3, 79 (2025).

51. Madeira, M. H., Boia, R., Santos, P. F., Ambrósio, A. F. & Santiago, A. R. Contribution of microglia-mediated neuroinflammation to retinal degenerative diseases. Mediators Inflamm. 2015, 673090 (2015).

52. Noailles, A., Maneu, V., Campello, L., Lax, P. & Cuenca, N. Systemic inflammation induced by lipopolysaccharide aggravates inherited retinal dystrophy. Cell Death Dis. 9, 350 (2018).

53. Shi, F.-D. & Yong, V. W. Neuroinflammation across neurological diseases. Science 388, eadx0043 (2025).

54. Makishima, H. et al. Germ line DDX41 mutations define a unique subtype of myeloid neoplasms. Blood 141, 534–549 (2023).

55. Weinreb, J. T. & Bowman, T. V. Clinical and mechanistic insights into the roles of DDX41 in haematological malignancies. FEBS Lett. 596, 2736–2745 (2022).

56. Baliakas, P. et al. How to manage patients with germline DDX41 variants: Recommendations from the Nordic working group on germline predisposition for myeloid neoplasms. HemaSphere 8, e145 (2024).

57. Rivolta, C. et al. RetiGene, a comprehensive gene atlas for inherited retinal diseases. Am. J. Hum. Genet. 112, 2253–2265 (2025).

58. Ajmal, M. et al. A missense mutation in the splicing factor gene DHX38 is associated with early-onset retinitis pigmentosa with macular coloboma. J. Med. Genet. 51, 444–448 (2014).

59. Krol, J. et al. A network comprising short and long noncoding RNAs and RNA helicase controls mouse retina architecture. Nat. Commun. 6, 7305 (2015).

60. Obuća, M., Cvačková, Z., Kubovčiak, J., Kolář, M. & Staněk, D. Retinitis pigmentosa-linked mutation in DHX38 modulates its splicing activity. PLOS ONE 17, e0265742 (2022).

61. Abramson, J. et al. Accurate structure prediction of biomolecular interactions with AlphaFold 3. Nature 630, 493–500 (2024).

62. Montpetit, B. et al. A conserved mechanism of DEAD-box ATPase activation by nucleoporins and InsP6 in mRNA export. Nature 472, 238–242 (2011).

63. Goddard, T. D. et al. UCSF ChimeraX: Meeting modern challenges in visualization and analysis. Protein Sci. Publ. Protein Soc. 27, 14–25 (2018).

64. Bankhead, P. et al. QuPath: Open source software for digital pathology image analysis. Sci. Rep. 7, 16878 (2017).

65. Fica, S. M., Oubridge, C., Wilkinson, M. E., Newman, A. J. & Nagai, K. A human postcatalytic spliceosome structure reveals essential roles of metazoan factors for exon ligation. Science 363, 710–714 (2019).

66. De Malglaive, F. et al. Pharmacological cAMP stimulation via prostaglandin receptors rescues ciliary defects in CEP290-deficient human and mouse models. Preprint at 10.1101/2023.10.06.561156 (2023).

67. Barny, I. et al. Basal exon skipping and nonsense-associated altered splicing allows bypassing complete CEP290 loss-of-function in individuals with unusually mild retinal disease. Hum. Mol. Genet. 27, 2689–2702 (2018).

68. Mutterer, J. & Zinck, E. Quick-and-clean article figures with FigureJ. J. Microsc. 252, 89–91 (2013).

69. Schindelin, J., et al. Fiji: an open-source platform for biological-image analysis. Nat. Methods 9, 676–682 (2012).

70. Liao, Y., Smyth, G. K. & Shi, W. featureCounts: an efficient general purpose program for assigning sequence reads to genomic features. Bioinforma. Oxf. Engl. 30, 923–930 (2014).

71. Robinson, M. D., McCarthy, D. J. & Smyth, G. K. edgeR: a Bioconductor package for differential expression analysis of digital gene expression data. Bioinforma. Oxf. Engl. 26, 139–140 (2010).

72. Singh, G. & Cooper, T. A. Minigene reporter for identification and analysis of cis elements and trans factors affecting pre-mRNA splicing. BioTechniques 41, 177–181 (2006).

