## Supplementary Data (Tables 1-3, Figures 1-4) for "Biallelic germline variants in the hematologic malignancy predisposition gene *DDX41* cause retinal dystrophy through dysregulation of retinal homeostasis"

### SUPPLEMENTARY TABLES

**Supplementary Table. 1 | Summary of *DDX41* variants identified in 13 affected individuals from 9 families.**

N.A.: not available; ACMG: American College of Medical Genetics and Genomics; M: mutation; PM: pathogenic moderate; PP: pathogenic supporting, PS: pathogenic strong. SpliceAI predicted probabilities (0-1; 0 = variant unlikely to affect splicing, 1 = likely splice-disrupting variant); AL: Acceptor Loss, DL: Donor Loss, AG: Acceptor Gain, DG: Donor Gain; *: according to the automated annotation in Alamut v1.13; $: according to gnomAD v2.1.1

| | P1 | P2 | P3 | P4 | P5 | P6 | P7 | P8 | P9 | P10 | P11 | P12 | P13 |
|---|---|---|---|---|---|---|---|---|---|---|---|---|---|
| Maternal variant NM_016222.4 NP_057306.2 | c.1187T>C p.Ile396Thr | c.1187T>C p.Ile396Thr | c.1015C>Tp.Arg339Cys | c.1187T>C p.Ile396Thr | c.1187T>C p.Ile396Thr | c.1187T>C p.Ile396Thr | c.1187T>C p.Ile396Thr | c.1015C>T p.Arg339Cys | c.1015C>T p.Arg339Cys | c.798G>A p.Ser266Ser Splicing event (deletion of exon 8 = p.leu216Glyfs*37) | c.655C>T p.Arg219Cys | c.962C>T p.Pro321Leu | c.962C>T p.Pro321Leu |
| Allele Frequency $ | 0.00001592 | 0.00001592 | 0.00003184 | 0.00001592 | 0.00001592 | 0.00001592 | 0.00001592 | 0.00003184 | 0.00003184 | 0.00002005 | N.A. | N.A. | N.A. |
| Splice AI | AL : 0.01<br>DL : 0.01<br>AG : 0.00<br>DG : 0.00 | AL : 0.01<br>DL : 0.01<br>AG : 0.00<br>DG : 0.00 | AL : 0.03<br>DL : 0.00<br>AG : 0.00<br>DG : 0.01 | AL : 0.01<br>DL : 0.01<br>AG : 0.00<br>DG : 0.00 | AL : 0.01<br>DL : 0.01<br>AG : 0.00<br>DG : 0.00 | AL : 0.01<br>DL : 0.01<br>AG : 0.00<br>DG : 0.00 | AL : 0.01<br>DL : 0.01<br>AG : 0.00<br>DG : 0.00 | AL : 0.03<br>DL : 0.00<br>AG : 0.00<br>DG : 0.01 | AL : 0.03<br>DL : 0.00<br>AG : 0.00<br>DG : 0.01 | AL : 0.32<br>DL : 0.37<br>AG : 0.03<br>DG : 0.20 | AL : 0.00<br>DL : 0.00<br>AG : 0.19<br>DG : 0.00 | AL : 0.00<br>DL : 0.00<br>AG : 0.02<br>DG : 0.00 | AL : 0.00<br>DL : 0.00<br>AG : 0.02<br>DG : 0.00 |
| ACMG class* | Likely Pathogenic (PM1, PP3, PP5) | Likely Pathogenic (PM1, PP3, PP5) | Uncertain Significance (score: 4 ; PM1, PM2) | Likely Pathogenic (PM1, PP3, PP5) | Likely Pathogenic (PM1, PP3, PP5) | Likely Pathogenic (PM1, PP3, PP5) | Likely Pathogenic (PM1, PP3, PP5) | Uncertain Significance (score: 4 ; PM1, PM2) | Uncertain Significance (score: 4 ; PM1, PM2) | Uncertain Significance (score: 1 ; PM2, BP4) | Uncertain Significance (score: 5; PM1, PM2, PP3) | Uncertain Significance (score: 5 ; PM1, PM2, PP3) | Uncertain Significance (score: 5 ; PM1, PM2, PP3) |
| Paternal Variant NM_016222.4 NP_057306.2 | c.1187T>C p.Ile396Thr | c.1187T>C p.Ile396Thr | c.1187T>C p.Ile396Thr | c.1187T>C p.Ile396Thr | c.1187T>C p.Ile396Thr | c.1187T>C p.Ile396Thr | c.305_306 delp.Lys102Argfs*32 | c.1015C>T p.Arg339Cys | c.1015C>T p.Arg339Cys | c.418_419insGTAGp.Asp140Glyfs*19 | c.655C>Tp.Arg219Cys | c.937G>C p.Gly313Arg | c.937G>C p.Gly313Arg |
| Allele Frequency $ | 0.00001592 | 0.00001592 | 0.00001592 | 0.00001592 | 0.00001592 | 0.00001592 | 0.00001591 | 0.00003184 | 0.00003184 | N.A. | N.A. | 0.000004000 | 0.000004000 |
| Splice AI | AL : 0.01<br>DL : 0.01<br>AG : 0.00<br>DG : 0.00 | AL : 0.01<br>DL : 0.01<br>AG : 0.00<br>DG : 0.00 | AL : 0.01<br>DL : 0.01<br>AG : 0.00<br>DG : 0.00 | AL : 0.01<br>DL : 0.01<br>AG : 0.00<br>DG : 0.00 | AL : 0.01<br>DL : 0.01<br>AG : 0.00<br>DG : 0.00 | AL : 0.01<br>DL : 0.01<br>AG : 0.00<br>DG : 0.00 | AL : 0.02<br>DL : 0.01<br>AG : 0.03<br>DG : 0.00 | AL : 0.03<br>DL : 0.00<br>AG : 0.00<br>DG : 0.01 | AL : 0.03<br>DL : 0.00<br>AG : 0.00<br>DG : 0.01 | AL : 0.09<br>DL : 0.03<br>AG : 0.00<br>DG : 0.00 | AL : 0.00<br>DL : 0.00<br>AG : 0.19<br>DG : 0.00 | AL : 0.02<br>DL : 0.00<br>AG : 0.00<br>DG : 0.00 | AL : 0.02<br>DL : 0.00<br>AG : 0.00<br>DG : 0.00 |
| ACMG class* | Likely Pathogenic (PM1, PM2, PP3, PP5) | Likely Pathogenic (PM1, PP3, PP5) | Likely Pathogenic (PM1, PP3, PP5) | Likely Pathogenic (PM1, PP3, PP5) | Likely Pathogenic (PM1, PP3, PP5) | Likely Pathogenic (PM1, PP3, PP5) | Pathogenic (score: 11; PVS1, PP5) | Uncertain Significance (score: 4 ; PM1, PM2) | Uncertain Significance (score: 4 ; PM1, PM2) | Pathogenic (score: 10; PVS1,) | Uncertain Significance (score: 5; PM1, PM2, PP3) | Uncertain Significance (score: 5; PM1, PM2, PP3) | Uncertain Significance (score: 5; PM1, PM2, PP3) |

**Supplementary Table. 2 Summary of clinical features of 13 affected individuals from 9 families with *DDX41* variants.**

Ophthalmological, neurological and skeletal findings in all affected individuals. +: symptoms present; −: symptoms absent; BE: both eyes; BPDCN: blastic plasmacytoid dendritic cell neoplasm; ERG: electroretinogram; EZ: ellipsoid zone; LE: left eye; LP: light perception; LRE: Left and right eyes; MEI: medico-educational institute; N.A.: not available; NLP: no light perception; OCT: optical coherence tomography; RE: right eye

| **Patient and Family** | **P1 F1** | **P2 F1** | **P3 F2** | **P4 F3** | **P5 F3** | **P6 F4** | **P7 F5** | **P8 F6** | **P9 F6** | **P10 F7** | **P11 F8** | **P12 F9** | **P13 F9** |
|---|---|---|---|---|---|---|---|---|---|---|---|---|---|
| Sex | Male | Male | Male | Female | Male | Female | Female | Female | Female | Male | Female | Female | Female |
| Ophthalmological features | | | | | | | | | | | | | |
| Age at diagnosis | 1-5yo | 1-5yo | 1-5yo | 1-5yo | 1-5yo | 1-5yo | 1-5yo | 11-15yo | 1-5yo | 1-5yo | 11-15yo | 6-10yo | 6-10yo |
| Nystagmus | N.A. | + | N.A. | + | + | + | + | + | + | N.A. | - | - | - |
| Photophobia | N.A. | + | + | + | + | + | - | N.A. | N.A. | N.A. | - | + | - |
| Night blindness | N.A. | - | + | N.A. | N.A. | N.A. | N.A. | N.A. | N.A. | + | + | - | - |
| BCVA | LP LRE | 1/10; LP LRE | 20/25 LRE | LP RE; hand movement LE | Hand movement LRE | LP LRE | N.A. | NLP LRE | LP LRE | 20/30 (-2) RE, 20/30 (-1) LE | LP RE, 2/60 LE | 20/50 LRE | 20/40 RE, 20/30 LE |
| Refractive error | N.A. | N.A | +1.50 -1.50 × 20° RE, +1.75 -1.00 × 180° LE | N.A. | N.A. | N.A. | N.A. | N.A. | N.A. | N.A. | −2.25 −1.00 × 12° RE, −6.75 −3.25 × 33° LE Keratoconus | 0.00 −3.00 × 15° RE, +1.00 −3.50 × 155° LE: | −0.25 −0.75 × 90° RE, −1.00 −1.00 × 90° LE (no cycloplegic) |
| ERG | Non-recordable | Non-recordable | Non-recordable | N.A. | Dominant cone dysfunction | N.A. | Generalized rod-cone dysfunction | N.A. | N.A. | Generalized rod-cone dysfunction | Severe rod-cone dysfunction | N.A. | N.A. |
| Fundus | N.A. | Peripheral pigment migration | Peripheral pigment migration | Bilateral, symmetrical chorioretinal atrophy with peripheral bone-like spicules and posterior/mid/far-atrophic areas | Bilateral, symmetrical chorioretinal atrophy with peripheral bone-like spicules and posterior/mid/far-atrophic areas | Diffuse pigment migration, narrow retinal vessels, pale optic disc | N.A. | Bilateral symmetrical chorioretinal atrophy with pigment migration, nummular mid/far-peripheral atrophy, pale optic discs, narrow vessels; posterior pole poorly seen (especially LE; media opacity | Bilateral symmetrical chorioretinal atrophy with pigment migration, nummular posterior/mid/far-peripheral atrophy, pale optic discs, narrow vessels, para-arterial sparing in mid-periphery | Bilateral blunted, narrow retinal vessels, mid-peripheral pigment migration, marked optic disc pallor with fibrosis in RE | Mid peripheral pigment mottling and pigment migration, narrow retinal vessels, pale optic disc | Normal | Normal |

| | | | | | | | | | | | | | |
|---|---|---|---|---|---|---|---|---|---|---|---|---|---|
| OCT | N.A. | N.A. | Global retinal thinning, normal retrofoveal EZ | Severe central macular thinnig (~150 µm LRE); inner retinal layers unsegmented (previously visible); outer retina lacks EZ | Very thin retina | N.A. | N.A. | Marked retinal disorganization with pigment migration LRE | Marked disorganization of the retina with pigment migration LRE | Central macular thickness: 213 µm RE, 212 µm LE | Generalized outer retinal disruption involving the fovea | Normal | Normal |
| Autofluorescence | N.A. | N.A. | N.A | N.A. | N.A. | N.A. | N.A. | N.A | Hypoautofluorescence in posterior pole and mid/far-periphery (overlapping bone spicules and nummular patches of atrophy) | Oval moderate hypoautofluorescence centered on macula, surrounded by hyperautofluorescence | Peripapillary hypoautofluorescence, central mottled hyperautofluorescence at the macula | N.A. | N.A. |
| Neurological features | | | | | | | | | | | | | |
| Neurological examination | Balance disorder; normal intellectual and psychomotor development | Normal intellectual and psychomotor development | Unremarkable | Unremarkable | Unremarkable | Cerebellitis | Cerebellar ataxia, developmental delay. Sentences not acquired but word associations, good interactions. | Ataxia, febrile seizures, psychomotor delay, mild dysarthria | Febrile seizures plus, psychomotor delay, mild dysarthria, ataxia | Cerebellar ataxia and atrophy, language delay, intellectual disability | Mild cognitive impairment | Moderate to severe psychomotor development delay | Mild to moderate psychomotor development delay |
| MRI observation | Vermis atrophy | Vermis atrophy | Infectious encephalopathy pattern | N.A. | N.A. | N.A. | Diffuse cerebellar atrophy | Vermis atrophy | Vermis atrophy | N.A. | N.A. | Mild ventricular enlargement | Mild ventricular enlargement |
| Skeletal features | | | | | | | | | | | | | |
| Morphological examination | Unremarkable | Unremarkable | Unremarkable | Unremarkable | Unremarkable | Unremarkable | Microcephaly (−2 to −3 SD; first sign) | Short stature, brachydactyly | Unremarkable | Microcephaly, short stature, clinodactyly and hypoplasic middle | Postaxial polydactyly (small extrafinger); small cutaneous lesion on finger side | Scoliosis | N.A. |

| | | | | | | | | | | | | | |
|---|---|---|---|---|---|---|---|---|---|---|---|---|---|
| | | | | | | | | | | phalana | | | |
| Hematological features | | | | | | | | | | | | | |
| | Unremarkable | Unremarkable | Unremarkable | Unremarkable | Unremarkable | Unremarkable | N.A. | N.A. | N.A. | N.A. | Anaemia on ferrous fumarate; Hb 114 g/L (120-150); low MCH & MCV | N.A. | Blastic plasmacytoid dendritic cell neoplasm. Now in remission. |
| Other | | | | | | | | | | | | | |
| | N.A. | N.A. | N.A. | N.A. | N.A. | N.A. | N.A. | N.A. | N.A. | Bladder Cancer | Hypothyroidism (on levothyroxine); hypophosphatemic rickets; delayed menarche; polycystic ovaries | N.A. | N.A. |

11

**Supplementary Table. 3 List of 8 differentially spliced genes (DSGs) and their splicing events in *Ddx41*$^{I396T/I396T}$ vs. wild-type littermates' retinas**

IJC: Inclusion Junction Counts, SJC: Skipping Junction Counts

| Gene.name | event | deltaPSI | KIKI_meanPSI | WT_meanPSI | KIKI.1_IJC | KIKI.2_IJC | KIKI.3_IJC | KIKI.4_IJC | KIKI.1_SJC | KIKI.2_SJC | KIKI.3_SJC | KIKI.4_SJC | WT.1_IJC | WT.2_IJC | WT.3_IJC | WT.4_IJC | WT.1_SJC | WT.2_SJC | WT.3_SJC | WT.4_SJC |
|---|---|---|---|---|---|---|---|---|---|---|---|---|---|---|---|---|---|---|---|---|
| Ppp4c | A3SS | -0,23 | 0,30 | 0,53 | 47 | 40 | 36 | 30 | 45 | 30 | 27 | 43 | 37 | 44 | 43 | 47 | 15 | 8 | 27 | 16 |
| Serf2 | SE | 0,22 | 0,75 | 0,53 | 88 | 87 | 79 | 55 | 6 | 5 | 8 | 3 | 73 | 57 | 69 | 67 | 17 | 7 | 16 | 12 |
| Rnf123 | RI | 0,25 | 0,62 | 0,38 | 156 | 181 | 151 | 184 | 29 | 16 | 24 | 15 | 209 | 181 | 188 | 183 | 70 | 58 | 64 | 62 |
| Laptm4a | MXE | -0,28 | 0,49 | 0,77 | 10 | 10 | 8 | 9 | 14 | 11 | 10 | 9 | 14 | 7 | 10 | 21 | 4 | 2 | 5 | 7 |
| Mrps17 | A5SS | 0,22 | 0,71 | 0,48 | 748 | 754 | 683 | 763 | 173 | 158 | 153 | 136 | 483 | 356 | 515 | 570 | 252 | 216 | 275 | 297 |
| Mrps17 | A5SS | 0,22 | 0,72 | 0,50 | 1036 | 991 | 956 | 1029 | 173 | 158 | 153 | 136 | 642 | 523 | 689 | 764 | 252 | 216 | 275 | 297 |
| Mrps17 | A5SS | 0,22 | 0,70 | 0,48 | 943 | 913 | 846 | 931 | 173 | 158 | 153 | 136 | 595 | 461 | 627 | 708 | 252 | 216 | 275 | 297 |
| Mrps17 | A5SS | 0,22 | 0,69 | 0,47 | 993 | 972 | 891 | 1019 | 173 | 158 | 153 | 136 | 658 | 506 | 683 | 764 | 252 | 216 | 275 | 297 |
| Mrps17 | RI | 0,21 | 0,51 | 0,30 | 1742 | 1707 | 1528 | 1824 | 173 | 158 | 153 | 136 | 1223 | 993 | 1144 | 1310 | 252 | 216 | 275 | 297 |
| Saxo2 | SE | -0,23 | 0,33 | 0,55 | 41 | 34 | 21 | 34 | 27 | 18 | 17 | 26 | 61 | 50 | 72 | 50 | 20 | 10 | 16 | 17 |
| Gas5 | MXE | -0,23 | 0,26 | 0,48 | 328 | 302 | 324 | 224 | 1103 | 1033 | 1106 | 1039 | 436 | 463 | 411 | 624 | 641 | 554 | 628 | 767 |
| Gas5 | MXE | -0,23 | 0,26 | 0,49 | 328 | 302 | 322 | 225 | 1103 | 1033 | 1106 | 1039 | 436 | 463 | 411 | 625 | 641 | 554 | 628 | 767 |
| Mrps17 | A5SS | 0,22 | 0,70 | 0,48 | 924 | 889 | 829 | 918 | 173 | 158 | 153 | 136 | 585 | 454 | 612 | 694 | 252 | 216 | 275 | 297 |
| Mrps17 | A5SS | 0,23 | 0,69 | 0,47 | 933 | 899 | 836 | 921 | 173 | 158 | 153 | 136 | 592 | 458 | 621 | 697 | 252 | 216 | 275 | 297 |
| Mrps17 | A5SS | 0,22 | 0,69 | 0,46 | 982 | 952 | 877 | 996 | 173 | 158 | 153 | 136 | 644 | 497 | 665 | 753 | 252 | 216 | 275 | 297 |
| Mrps17 | A5SS | 0,22 | 0,68 | 0,45 | 985 | 956 | 890 | 1006 | 173 | 158 | 153 | 136 | 647 | 505 | 673 | 752 | 252 | 216 | 275 | 297 |
| Gm57315 | A3SS | 0,32 | 0,66 | 0,34 | 8 | 12 | 7 | 10 | 3 | 6 | 4 | 4 | 11 | 5 | 13 | 12 | 22 | 16 | 16 | 14 |

### SUPPLEMENTARY FIGURES

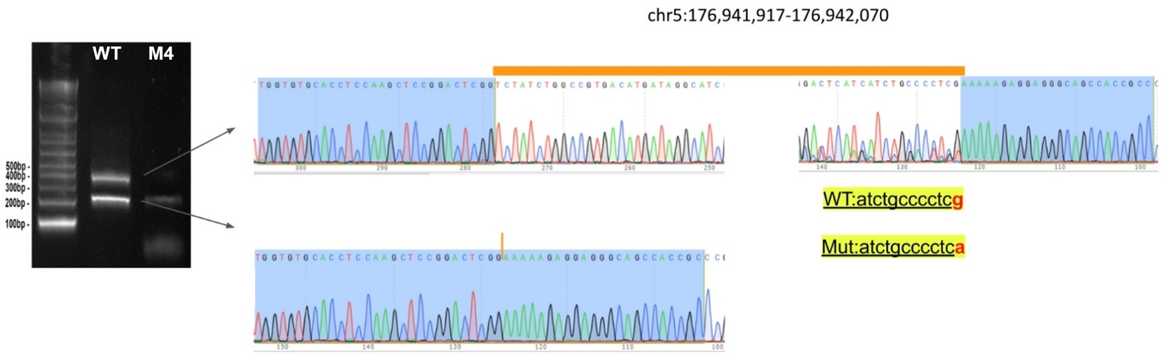


**Supplementary Fig.1  Minigene assay with M4 confirmed the modification of splicing.** Minigene assay showing that the M4 variant (c.798G>A, p.Ser266=) alters pre-mRNA splicing by skipping exon 8 (p.Leu216Glyfs*37). In cells transfected with the M4 construct, RT-PCR analysis revealed an abnormal splice product compared with the wild-type minigene, confirming that M4 modifies splicing. This variant is positioned in GRCh38.

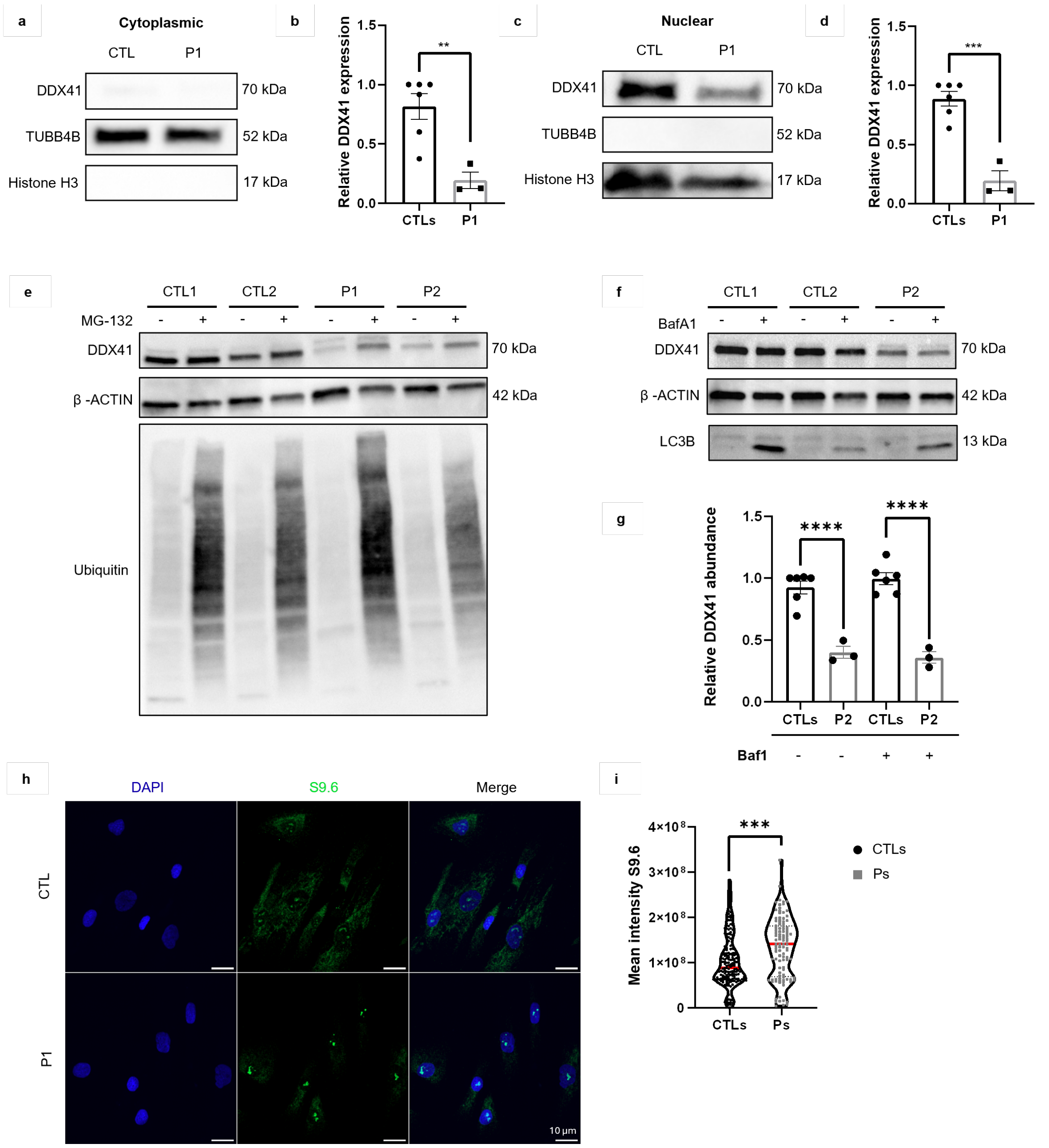


**Supplementary Fig. 2 Analysis of DDX41 expression and subcellular localization in control and patient fibroblasts.**

(a, b) DDX41 levels in cytoplasmic fractions from controls (CTLs) and patient (P1) fibroblasts. (a) Representative Western blot; TUBB4B was used as a loading control, and molecular weight markers are indicated on the right. (b) Quantification of cytoplasmic DDX41 normalized to TUBB4B, showing reduced cytoplasmic localization in patient fibroblasts compared with controls. Data represent mean ± SEM ($n = 3$). ***, $p = 0.0003$ (unpaired two-tailed Student t-test). (c, d) DDX41 levels in nuclear fractions from controls (CTLs) and patient (P2) fibroblasts. (c) Representative Western blot; histone H3 was used as a loading control. Molecular weight markers are indicated on the right. (d) Quantification of nuclear DDX41 normalized to histone H3, showing decreased nuclear localization in patient fibroblasts compared with controls. Data represent mean ± SEM ($n = 3$). **: $p = 0.007$ (unpaired two-tailed Student t-test). (e) Western blot analysis of DDX41 expression in control (CTL1 and CTL2) and patient (P1 and P2) fibroblasts following treatment with 10 µM MG-132 for 18 h. β-actin serves as a loading control, and ubiquitin is shown as a positive control for proteasome inhibition. (f, g) Effect of lysosomal inhibition on DDX41 levels. (f) Western blot analysis of DDX41 expression in control (CTL1 and CTL2) and patient (P2) fibroblasts after treatment with bafilomycin A1 (BafA1); β-actin was used as a loading control. (g) Quantification of DDX41 protein levels normalized to β-actin, comparing untreated and BafA1-treated samples. Data represent mean ± SEM ($n = 3$). ****, $p < 0.0001$ (two-way ANOVA with Tukey post hoc test) (h, i) R-loop accumulation in control and patient fibroblasts. (h) Immunofluorescence detection of R-loops using the S9.6 antibody; increased nuclear S9.6 signal intensity was observed in patient cells. Scale bars, 10 μm. (i) Quantification of nuclear R-loop levels from S9.6 staining. Prior to statistical analysis, data were subjected to outlier detection using the ROUT test ($Q = 1\%$). Remaining data are shown as mean ± SEM from three independent experiments. ***, $p = 0.0002$ (unpaired two-tailed Student t-test).

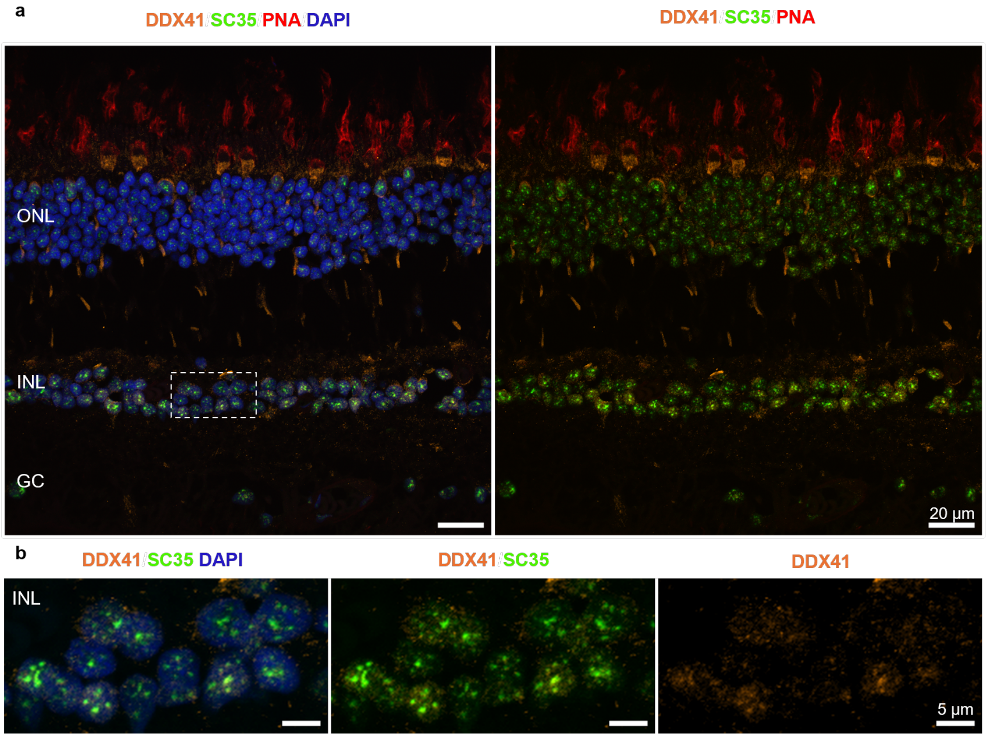


**Supplementary Fig.3 Immunostaining of DDX41 in the human retina.**

(a) Immunofluorescence staining of transverse sections from paraffin-embedded human retina reveals the presence of DDX41 (orange), predominantly within the inner nuclear layer (INL) of this tissue. DDX41 partially colocalizes with nuclear speckles marked by SC35 (green). Cones are labeled with PNA (peanut agglutinin, red), DAPI (4′,6-diamidino-2-phenylindole, blue) stains nuclei. ONL, outer nuclear layer; GC, ganglion cell layer. Scale bars: 20 µm. (b) Higher magnification of a selected retinal region, focusing on the INL, highlights the nuclear distribution of DDX41 and its partial colocalization with SC35-marked nuclear speckles. Scale bars: 5 µm.

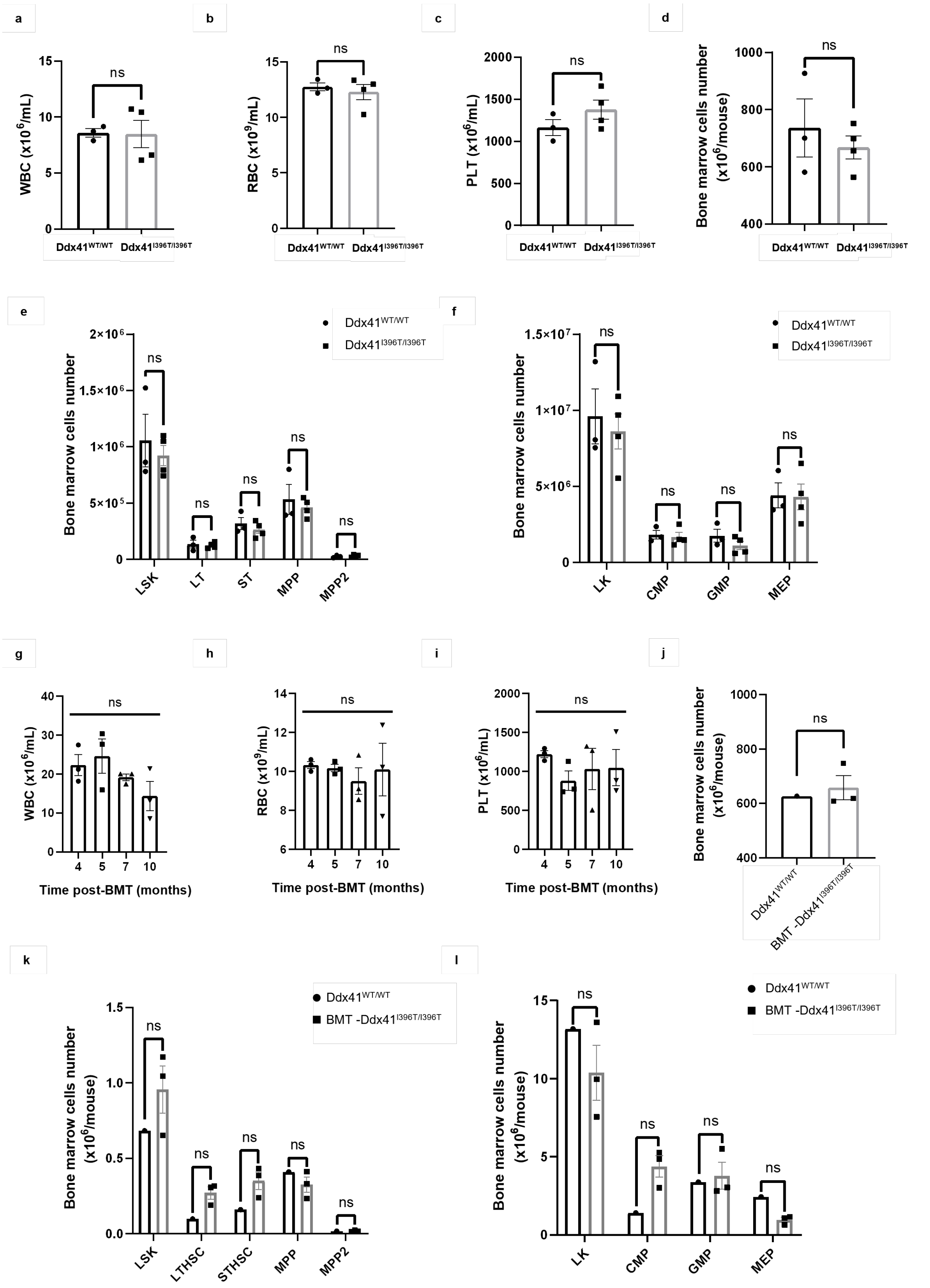


**Supplementary Fig. 4 Hematological evaluation of *Ddx41*$^{I396T/I396T}$ mice and bone marrow-transplanted (BMT) *Ddx41*$^{I396T/I396T}$ mice.**

(a-c) Peripheral blood counts in wildtype (WT) and *Ddx41*$^{I396T/I396T}$ mice. (a) white blood cells (WBC); (b) red blood cells (RBC) and (c) platelets (PLT). Data are mean ± SEM; n.s., not significant (unpaired two-tailed Student t-test). (d-f) Bone marrow hematopoietic compartments analyzed by flow cytometry in WT and $Ddx41^{I396T/I396T}$ mice. (d) Total bone marrow cells (n.s., not significant; unpaired two-tailed Student t-test), (e) Total Lin$^-$ Sca-1$^+$ c-Kit$^+$ (LSK) and LSK-subpopulations (long-term hematopoietic stem cells (LT), short-term hematopoietic stem cells (ST), multipotent progenitors (MPP) and multipotent progenitor 2 (MPP2)). (f) Total Lin$^-$ c-Kit$^+$ (LK) and LK subpopulations (common myeloid progenitors (CMP); granulocyte-macrophage progenitors (GMP) and megakaryocyte-erythroid progenitors (MEP)). Data are mean ± SEM; n.s., not significant (two-way ANOVA with Sidak correction for multiple comparisons) (g-i) Peripheral blood counts in BMT-$Ddx41^{I396T/I396T}$ mice at 4, 5, 7, and 10 months post-transplantation. (g) WBC; (h) RBC; (i) PLT. Data are mean ± SEM; n.s., not significant (unpaired two-tailed Student t-test). **(j-l)** Bone marrow hematopoietic compartments in WT and BMT-$Ddx41^{I396T/I396T}$ mice at 10 months post-transplantation. (j) Total bone marrow cells (n.s., not significant; unpaired two-tailed Student t-test), (k) Total LSK and LSK-subpopulations (LT, ST, MPP, MMP2) (l) Total LK and LK subpopulations (CMP, GMP, MEP). Data are mean ± SEM; n.s., not significant (two-way ANOVA with Sidak correction for multiple comparisons).

#### SUPPLEMENTARY METHODS

##### *Minigene assay*

A minigene assay was performed to analyze the splicing effect[72]. The fragment used for minigene assay was chr5:177,514,848-177,515,177 (hg38). PCR was performed on normal and patient, and products were cloned into the RHCglo vector. Splicing effects were analyzed by transfecting HEK293 cells and performing RT-PCR. Sanger sequencing was performed for purified gel products. The primer sequences for RT-PCR are: Rhc-RT-F-CATTCACCACATTGGTGTGC and Rhc-RT-R- GTGTCACATGGAGCTTTGCA.

##### *Nuclear and Cytoplasmic Protein Extraction and Western Blot Analysis*

Primary dermal fibroblasts from two independent controls and patient P1 were harvested by trypsinization, washed once with ice-cold PBS, and subjected to differential extraction on ice to isolate cytoplasmic and nuclear fractions. Cell pellets were resuspended in hypotonic buffer (10 mM HEPES pH 7.9, 0.5 mM DTT, protease/phosphatase inhibitors), incubated on ice for 5 min, vortexed briefly after adding 0.4% NP-40, and centrifuged at 800 × g for 2 min at 4°C; supernatants were collected as cytoplasmic fractions. Nuclear pellets were washed three times with detergent-free hypotonic buffer, then resuspended in nuclear extraction buffer (20 mM Tris-HCl pH 7.9, 138 mM NaCl, 2.7 mM KCl, 5 mM EDTA, 20 mM NaF, 1% NP-40, 5% glycerol, protease/phosphatase inhibitors) and vortexed three times for 10 s each.

Western blot analysis was performed as described in the Materials and Methods section. Fraction purity was confirmed using rabbit polyclonal anti-Histone H3 (1:1400, ab1791, Abcam) for nuclear and mouse monoclonal anti-TUBB4B (1:6700, H00010383-M02, Abnova) for cytoplasmic markers. DDX41 was detected with primary and HRP-conjugated secondary antibodies followed by chemiluminescent detection.

##### *Bafilomycin A1 (BAF A1) Treatment and Western Blot Analysis*

Primary dermal fibroblasts from two independent controls and patient P2 were treated with Bafilomycin A1 (BAF A1; Sigma-Aldrich) at 10 nM for 24 h to inhibit autophagic flux via vacuolar H+-ATPase blockade, prepared in DMSO and diluted into culture medium, with control cells receiving equivalent DMSO vehicle.

Western blot analysis was performed as described in the Materials and Methods section. Treatment efficacy was verified using rabbit polyclonal anti-LC3B (1:500, 18725-1-AP, Proteintech) for autophagy. DDX41 was detected with primary and HRP-conjugated secondary antibodies followed by chemiluminescent detection.

##### *Immunostaining in the Human Retina*

Human retinal tissue obtained post-enucleation was fixed, paraffin-embedded, and sectioned. Sections were deparaffinized in xylene, rehydrated through graded ethanol, and subjected to antigen retrieval in 0.01 M citrate buffer (pH 6.0) at 98°C for 15 min. Sections were then permeabilized (0.2% fish skin gelatin, 0.25% Triton X-100 in 1× PBS), blocked for 1 h in permeabilization buffer with 5% normal goat serum, and incubated overnight with primary antibodies: anti-DDX41 (Invitrogen, PA5-109625, 1:500), anti-SC35 (Sigma-Aldrich, S4045, 1:200), and anti-GS (Merck, MAB302, 1:1000).

Secondary antibodies were Alexa Fluor 488 F(ab')$_2$ goat anti-rabbit IgG (Invitrogen, A-11070, 1:500) and Alexa Fluor 647 F(ab')$_2$ goat anti-mouse IgG (Invitrogen, A-21237, 1:500), with DAPI (Thermo Fisher, D1306, 1:500) for nuclei, WGA (Vector Labs, RL-1022-10, 1:1000) for rods, and PNA (Invitrogen, L32458, 1:1000) for cones. Staining specificity was verified by secondary-only controls. Sections were mounted in ProLong Gold Antifade Mountant (Thermo Fisher) with #1.5 coverslips and imaged using an Olympus confocal microscope (63× oil objective).

***Blood Count Analysis***

Peripheral Blood was collected from WT and mutant mice from the submandibular venous plexus into EDTA-coated tubes (BD #365975). Blood counts were performed using an automated hematology analyzer (MS9; Schloessing Melet, Cergy-Pontoise, France).

***Bone Marrow Transplantation and Analysis***

Bone marrow cells were isolated by spin-through, filtered through 70 μm strainers (Corning), and resuspended in PBS + 2% FBS. Single-cell suspensions underwent flow cytometry at Saint-Louis Hospital platform, with remaining cells transplanted into Ly5.1 recipient mice (10 × $10^6$ cells/mouse); recipients were monitored for up to 10 months.

Hematopoietic analysis used Zombie UV™ Fixable Viability Dye (BioLegend #423107; 15 min RT), followed by biotin-conjugated Lineage cocktail (BioLegend #133307; 30 min ice), then fluorochrome-conjugated antibodies: PerCP/Cy5.5 anti-CD117 (BioLegend #105824), BV510 anti-Sca-1 (BioLegend #108129), APC/Cy7 streptavidin (BioLegend #405208), BV711 anti-CD48 (BioLegend #103439), BV786 anti-CD16/32 (BD #740851), AF647 anti-CD34 (BD #560230), PE/Cy7 anti-CD150 (BioLegend #115914) (30 min 4°C dark). Cells were washed, acquired on Fortessa, and analyzed with FlowJo v10.
